# Genome-wide association studies in a large Korean cohort identify novel quantitative trait loci for 36 traits and illuminate their genetic architectures

**DOI:** 10.1101/2024.05.17.24307550

**Authors:** Yon Ho Jee, Ying Wang, Keum Ji Jung, Ji-Young Lee, Heejin Kimm, Rui Duan, Alkes L. Price, Alicia R. Martin, Peter Kraft

## Abstract

Genome-wide association studies (GWAS) have been predominantly conducted in populations of European ancestry, limiting opportunities for biological discovery in diverse populations. We report GWAS findings from 153,950 individuals across 36 quantitative traits in the Korean Cancer Prevention Study-II (KCPS2) Biobank. We discovered 301 novel genetic loci in KCPS2, including an association between thyroid-stimulating hormone and *CD36*. Meta-analysis with the Korean Genome and Epidemiology Study, Biobank Japan, Taiwan Biobank, and UK Biobank identified 4,588 loci that were not significant in any contributing GWAS. We describe differences in genetic architectures across these East Asian and European samples. We also highlight East Asian specific associations, including a known pleiotropic missense variant in *ALDH2*, which fine-mapping identified as a likely causal variant for a diverse set of traits. Our findings provide insights into the genetic architecture of complex traits in East Asian populations and highlight how broadening the population diversity of GWAS samples can aid discovery.

## Introduction

Large-scale biobanks integrating genomic and electronic health record data enable genome-wide association studies (GWAS) to identify numerous genetic associations and provide insights into the biological mechanisms of human complex traits and diseases.^1,2^ In turn, the combined effects of these genetic markers can be summarized as polygenic risk score (PRS) to estimate individuals’ genetic predispositions for complex diseases, which have successfully identified individuals with a high risk of disease.^3,4^ However, current genetic discovery efforts heavily underrepresent non-European populations globally and thus limit further discoveries of variants that are rare or absent in European (EUR) populations but common in other ancestry groups.^5^ Furthermore, this genomic research imbalance could lead to health disparities if the genomic discoveries benefit only European ancestry individuals in clinical practice.^6^

Early efforts toward diversifying GWAS in East Asian (EAS) populations, including Biobank Japan (BBJ),^7–9^ Korean Genome and Epidemiology Study (KoGES),^10–13^ and China Kadoorie Biobank,^14^ Taiwan Biobank (TWB)^15^ have made significant contributions and facilitated various genetic studies in these populations. Despite these efforts, the representation of EAS groups in genetic research remains low, compared to European groups (e.g., UK Biobank [UKB]^16^, FinnGen^17^, HUNT^18^, and deCODE^19^). For example, one of the most extensively used resources is UKB, which includes approximately 500,000 British individuals with deep phenotyping and genomic data.^20^ According to the GWAS Diversity Monitor,^21^ over 90% of total GWAS participants are from European-ancestry samples, while only 4% of participants are of Asian origin despite making up 59% of the global population. The inclusion of additional EAS biobanks is warranted to empower genetic discovery and elucidate the genetic architecture of complex traits and diseases within East Asia.

Here we conducted GWAS for 36 quantitative traits from 153,950 individuals in the Korean Cancer Prevention Study-II (KCPS-II) Biobank^22^, a prospective cohort study of the Korean population with genomic data and a wide range of measured phenotypes. Following the GWAS in KCPS2, we meta-analyzed 21 traits across KCPS2, KoGES, BBJ, TWB, and UKB to identify significant loci across East Asian and European ancestry populations. We compared the genetic architectures of these traits across populations leveraging GWAS summary statistics from KCPS2, KoGES, BBJ, TWB, and UKB. Lastly, we pinpointed putatively causal variants through fine-mapping and conducted colocalization to understand the biological mechanisms underlying these traits.

## Results

A total of 153,950 participants were genotyped, including 64,812 participants on the GSA-chip array and 89,138 participants on the Korean-chip array in this study. We subsequently conducted genotype quality control (QC) and imputation. Figure 1 provides an overview of the KCPS2 samples, the traits examined, their abbreviations, and the analyses conducted in this study (Table S1). We analyzed 36 quantitative traits including 4 anthropometric traits, 7 metabolic biomarkers, 5 liver function enzymes, 1 thyroid hormone, 1 tumor marker, 3 kidney function traits, 10 hematological traits, 2 cardiovascular traits, and 3 lifestyle factors.

**Figure 1.**
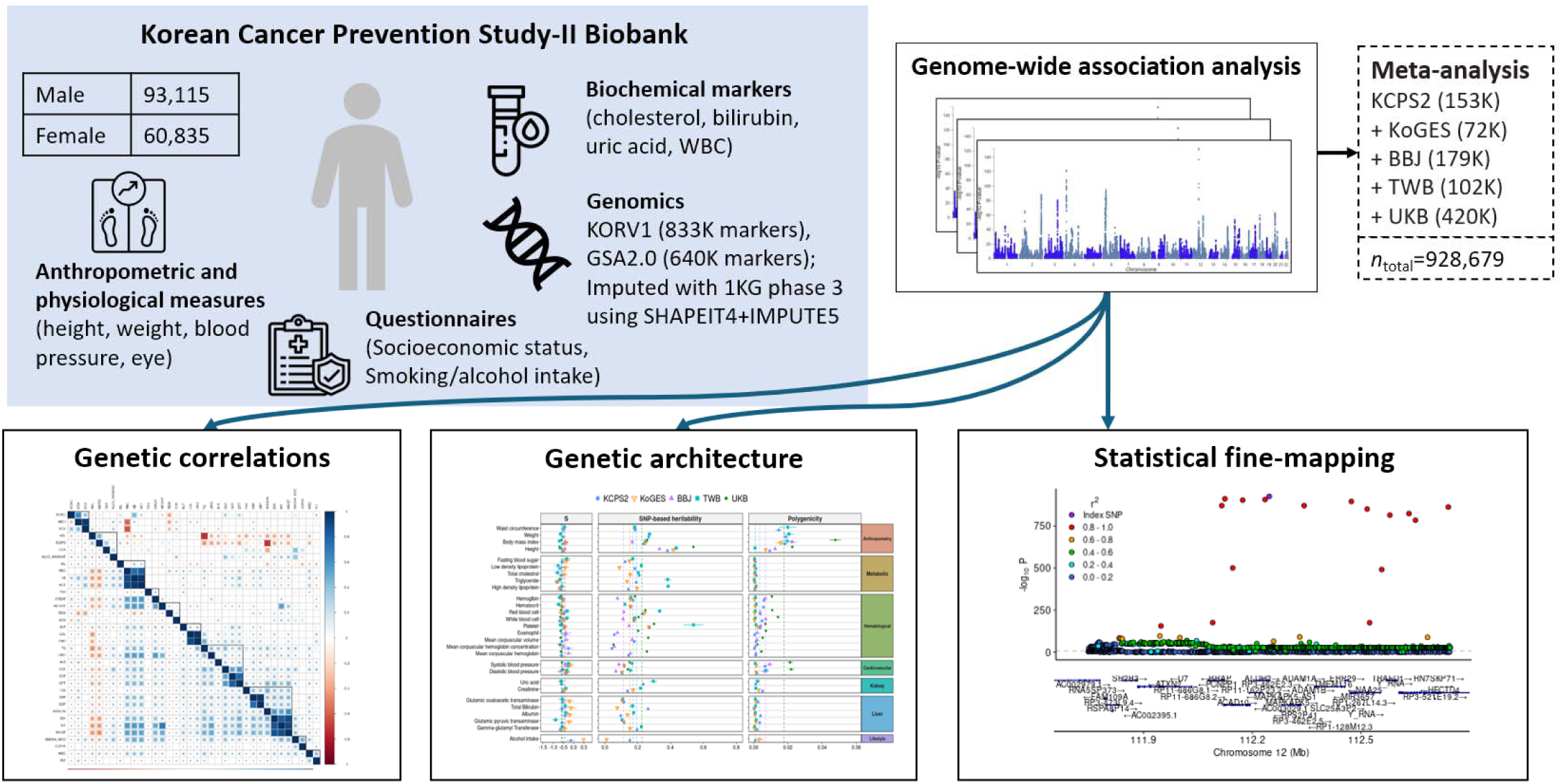
| Overview of the Korean Cancer Prevention Study-II Biobank and analysis. Detailed descriptions of the 36 quantitative traits examined in this study are shown in Table S1. After QC, the data were phased using SHAPEIT4^23^ and imputed using IMPUTE5^24^ with 1000 Genomes Project Phase 3 data.

### GWAS of KCPS2 and pleiotropy analysis

We conducted GWAS of 36 human quantitative traits in the KCPS2 Biobank (n=153,950). We used a linear mixed model implemented in SAIGE^25^ for association testing to maximize statistical power and included age, sex, 10 principal components (PCs), and SNP array as covariates. None of the GWAS exhibited striking systematic inflation in test statistics indicative of population stratification or other artifact (median λ_GC_ 1.23, median S-LDSC intercept 1.04) (Table S1). Using S-LDSC with the baseline-LD model, we estimated the SNP-based heritability for each trait (Table S1), which ranged from 0.034 (alcohol intake) to 0.347 (height).

Our analysis discovered 2,962 independent genome-wide significant loci (median 68, range 1-428 loci; 2,631 unique loci) across 36 traits using the 1000 Genome phase 3 EAS samples as the LD reference (Table S2). Among these, 301 loci (median 6, range 0-32 loci) were not reported in previous GWAS^26^ related to the corresponding trait using Experimental Factor Ontology (EFO) term (Figure 2a, Table S3), with the greatest fraction of novel loci (novelty rate) for carcinoembryonic antigen [CEA, 10/16 (63%) novel loci], followed by thyroid-stimulating hormone [TSH, 32/100 (32%) novel loci]. Across 21 traits available in all five biobanks, 36% of the novel loci replicated in a meta-analysis of KoGES, BBJ and TWB (Table S12). The novel loci tend to be more common in KCPS2 than in 1000 Genome phase 3 EUR samples (median KCPS2 minor allele frequencies [MAF]: 0.207 vs. median EUR MAF: 0.118; paired t test *P* = 2.2e-16 vs. median EAS MAF: 0.202; paired t-test P = 3.8e-4). (Figure S1). We also identified widespread pleiotropy: 4,960 gene regions contained variants associated with one or more traits (mean 2.3 traits, range 1-27). For example, out of 36 traits, variants near *ALDH2* were associated with 26 traits, including blood pressure and liver enzyme values (Figure 2b, Table S4).

**Figure 2.**
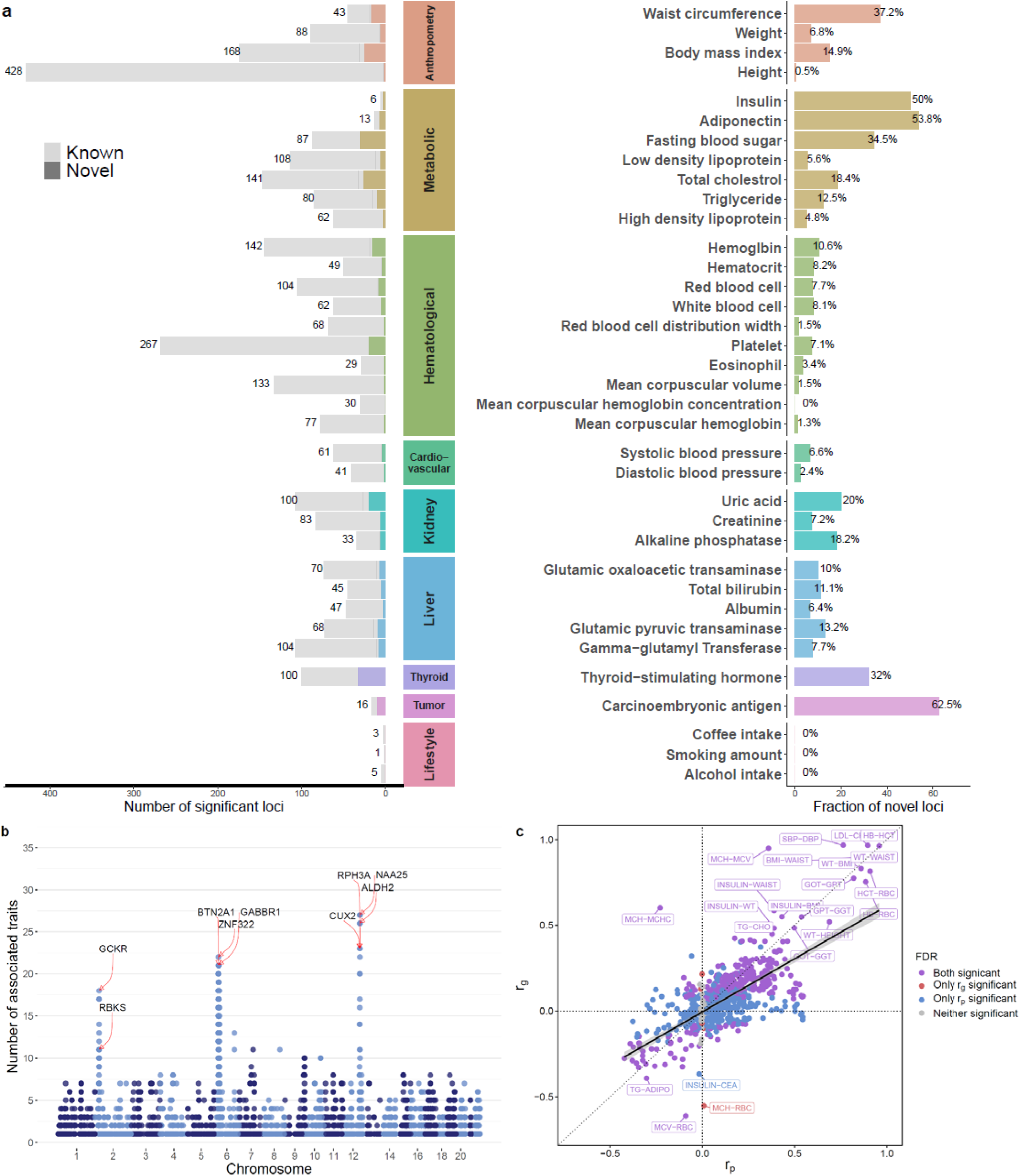
| GWAS results for 36 quantitative traits in the Korean Cancer Prevention Biobank-II (KCPS2). (a) Number of known and novel variants identified in KCPS2 compared to the Open Target Genetics^27^ using EFO terms (Table S2-S3). **(b)** A summary of genome-wide significant loci associated with the 36 traits in KCPS2. Each locus was mapped to a gene using FUMA^28^ with a 1000 Genome Phase 3 East Asian reference panel. We then counted the number of associated traits (out of 36 traits) per gene (Table S4). **(c)** Comparisons of pairwise genetic correlations (rg) between phenotypic correlations (rp) for the 36 traits in KCPS2. rg was estimated using bivariate LDSC based on association test statistics from linear regression. Significant rg and rp after false discovery rate (FDR<0.05) correction is indicated by purple if both rg and rp were significant, red if only rg was significant, blue if only rp was significant, and gray if neither was significant. The black solid line was estimated by spline smoothing from a linear regression model. The complete set of rg and rp is available in Table S5.

### Genetic and phenotypic correlations between the 36 traits in KCPS2

By estimating pairwise genetic correlations (r_g_) between traits, we identified clusters of highly genetically correlated traits, including cardiometabolic risk factors (e.g., fasting blood sugar [FBS], systolic blood pressure [SBP], diastolic blood pressure [DBP], insulin, body mass index [BMI], weight, and waist circumference) and liver enzyme traits (e.g., albumin, glutamic oxaloacetic transaminase [GOT], glutamic pyruvic transaminase [GPT], and gamma-glutamyl transferase [GGT]) (Table S5, Figure 2c, Figure S2). The slope of the relationship between pairwise genetic correlations and phenotypic correlations (r_p_) for 36 traits was 0.634. We identified significantly negative genetic and phenotypic correlations of high-density lipoprotein [HDL] cholesterol and adiponectin with the majority of cardiometabolic risk factors including FBS, insulin, and BMI (mean cardiometabolic traits r_g_=-0.24, r_p_=-0.25 for HDL; r_g_=-0.27, r_p_=-0.27 for adiponectin). These findings are consistent with a known cardioprotective role of HDL^29^ and beneficial effects of adiponectin on obesity-associated metabolic and vascular disorders.^30,31^ In contrast, the genetic correlations between bilirubin and a number of cardiometabolic risk factors (mean cardiometabolic traits [FBS, insulin, BMI, waist circumference] rg=-0.09), low-density lipoprotein cholesterol (r_g_=-0.13 [FDR=0.012]), and WBC (r_g_=-0.11 [FDR=0.001]) were significantly negative, although the phenotypic correlations were significantly positive (e.g., mean correlation between bilirubin and cardiometabolic traits r_p_=0.04). Bilirubin levels have been shown to be inversely correlated with cardiovascular disease risk by inhibiting cholesterol synthesis and modulating the immune system,^32,33^ which is supported by our genetic correlations results. Liver enzyme values such as GGT were positively associated with alcohol consumption both genetically and phenotypically (r_g_=0.31 [FDR=0.0001], r_p_=0.33 [FDR<0.0001]), consistent with a previous Mendelian randomization (MR) study.^34^ Similarly, smoking, alcohol consumption, and hemoglobin showed significantly positive genetic and phenotypic correlations with a number of cardiometabolic risk factors (mean cardiometabolic traits r_g_=0.18, r_p_=0.22 for smoking; r_g_=0.14, rp=0.16 for alcohol consumption; r_g_=0.14, r_p_=0.28 for hemoglobin). For hemoglobin, previous MR showed evidence for lower hemoglobin levels being associated with lower BMI, better glucose tolerance and other metabolic profiles, lower inflammatory load, and blood pressure.^35^

### Meta-analysis of 21 traits across KCPS2, KoGES, BBJ, TWB, and UKB

We meta-analyzed 21 traits across KCPS2 (153K), KoGES (72K), BBJ (179K), TWB (102K), and UKB (420K) and discovered a total of 12,224 loci associated with the 21 traits, among which 4,588 were not genome-wide significant in any of the other four contributing GWAS (Figure 3a, Figure S3, Table S6-S7). The median MAF in KCPS2 for the lead variants at the loci which were only significant in the meta-analysis but not significant in the other individual GWAS, was lower than the MAF in KCPS2 for the lead variants at the loci that were only significant in the KCPS2 (median MAF 0.29 versus 0.31, respectively) (Figure 3b).

**Figure 3.**
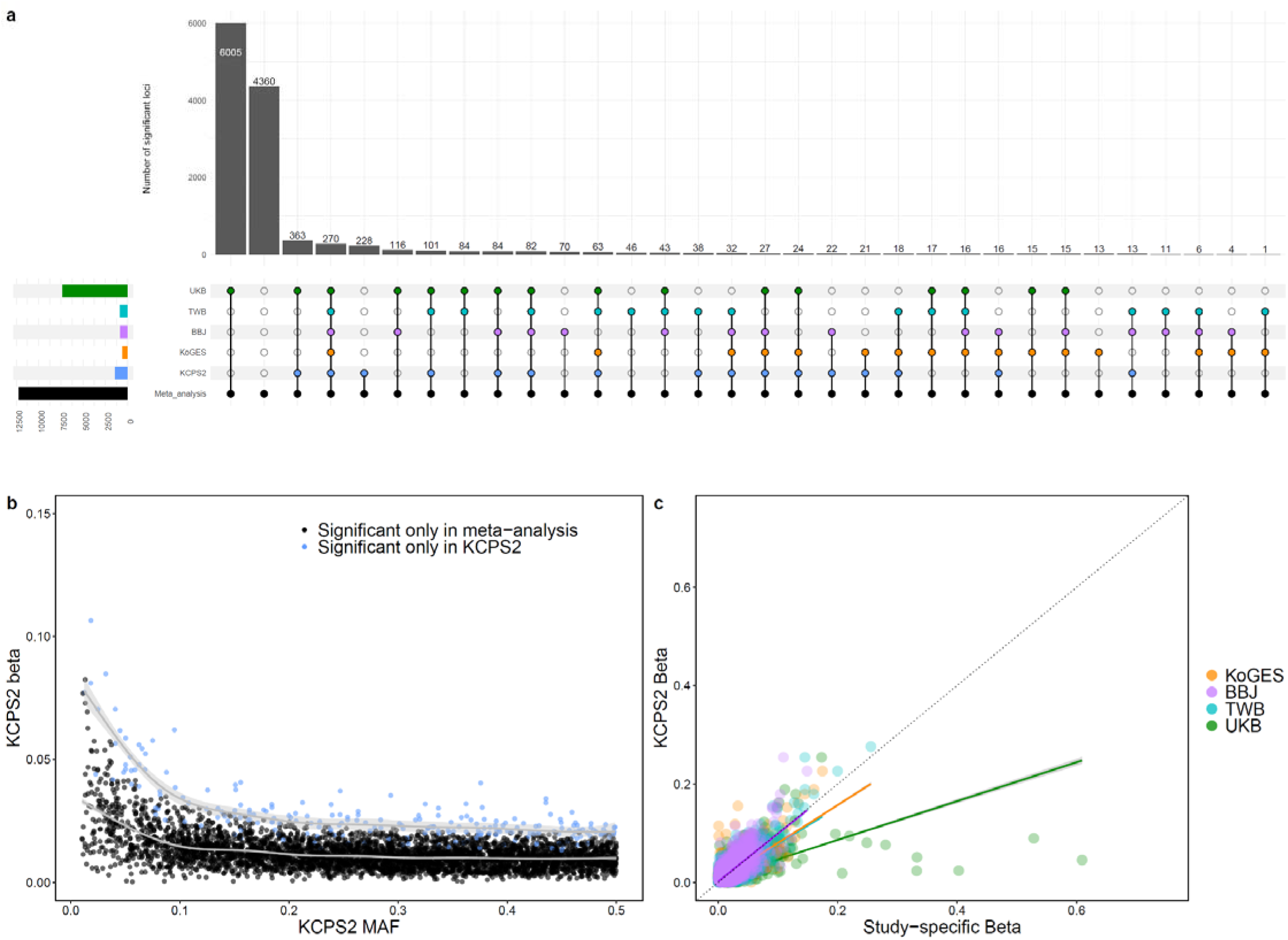
| Meta-analysis of 21 traits across KCPS2, KoGES, BBJ, TWB, and UKB. **(a)** Genome-wide significant loci identified in the meta-analysis, Color of dots indicate significance in meta-analysis (black), KCPS2 (blue), KoGES (orange), BBJ (purple), TWB (light blue), and UKB (green). Multiple dots in a bar represent simultaneous significance in multiple cohorts. **(b)** Comparisons of allele frequency and effect sizes in KCPS2 for the genome-wide significant variants discovered only in KCPS2 (blue) versus those identified only in the meta-analysis (black). **(c)** Comparisons of effect sizes in KCPS2 and study-specific effect sizes for the lead variants at the 12,224 meta-analysis genome-wide significant loci. The solid lines were estimated by spline smoothing from generalized additive model (b) or linear regression model (c). Full meta-analysis results are shown in Table S6-7.

We compared effect sizes from KCPS2 to effect sizes by study for the lead variants at the 12,224 genome-wide significant loci from meta-analysis (Figure 3c). The correlations between KCPS2 effect sizes and those in other East Asian biobanks are similar (KCPS2-KoGES: 0.890, KCPS2-BBJ: 0.887, KCPS2-TWB: 0.893), with smaller correlation with UKB (KCPS2-UKB: 0.765). To further explore novel associations, we compared effect sizes from the meta-analysis to MAF by study (Figure S4). There was an inverse relationship between MAF and effect size, due in part to the restriction to genome-wide significant variants. The lead variants from genome-wide significant loci identified in the meta-analysis had in general similar study-specific MAF in East Asian populations (KCPS2 [median MAF=0.27], KoGES [median MAF=0.27], BBJ [median MAF=0.27], and TWB [median MAF=0.28]) compared to European ancestry populations in UKB (median MAF=0.26).

### Genetic architecture compared between KCPS2, KoGES, BBJ, TWB, and UKB

We investigated the genetic architecture in KCPS2 (Figure S5) and compared it with KoGES, BBJ, TWB, and UKB across seven trait categories (Figure 4a, Table S8). The S parameters linking MAF and effect sizes were similar across the biobanks (median S=-0.51, range -0.80, -0.03), suggesting a pervasive action of negative selection on the trait-associated variants.^36^ The SNP-heritability estimates (*h^2^_g_*) varied widely across different biobanks and categories. For example, compared to BBJ, KCPS2 has higher heritability estimates for anthropometry (median *h^2^_g_* =0.31 vs. 0.25), cardiovascular (median *h^2^_g_* =0.11 vs. 0.08), and hematological traits (median lil2g=0.18 vs. 0.11). For hematological traits, UKB has the largest heritability estimates (median *h^2^_g_* =0.23), with the exception of platelet (*h^2^_g_* =0.54) and RBC (*h^2^_g_* =0.33) being the largest in TWB. Compared to TWB, KCPS2 has lower heritability estimates for metabolic (median *h^2^_g_* =0.18 vs. 0.22), liver (median *h^2^_g_* =0.12 vs. 0.17), and kidney traits (median *h^2^_g_* =0.19 vs. 0.26). However, these differences were not statistically significant (Wilcoxon signed-rank test p > 0.05), likely due to the limited number of traits being compared (number of paired comparisons range from 1 to 8) (Table S14). Overall, we observed high correlations of heritability in KCPS2 with the other biobank (KCPS2-KoGES: Pearson correlation r=0.70, P=4e-04; KCPS2-BBJ: r=0.86, P=0.00033; KCPS2-TWB: r=0.80, P=5.2e-06; and KCPS2-UKB: r=0.90, P=7.7e-05) (Figure 4b-e). We note that our TWB heritability estimates using SbayesS and unmatched LD were higher than those reported by Chen et al., (2023)^15^ using LDSC and in-sample LD, especially for metabolic and hematological traits (Figure S6a), which led to a low correlation between the heritability estimates of KCPS2 and TWB (r=0.64, 95% CI: 0.31-0.83, P=9.62e-04) (Figure S6b). When we used the heritability estimates reported by Chen and colleagues, the correlation in heritability between KCPS2 and TWB improved (r=0.80, 95% CI: 0.58-0.91) (Figure 4d). The most polygenic traits (weight and BMI) had about 2% SNPs with nonzero effects, whereas the least polygenic traits (coffee intake, bilirubin, and MCHC) were affected by about 0.02-0.03% common SNPs in KCPS2. The median polygenicity estimates for the 8 traits available in all five studies were largest in UKB (median π=0.02), followed by BBJ (median π=0.007), KCPS2 (median π=0.005), KoGES (median π=0.002), and TWB (median π=0.001), which in general follows the same order as the sample sizes of the biobanks. Nevertheless, the genetic correlation (r_g_) estimates within EAS were close to 1 (KCPS2-KoGES median r_g_=0.997, KCPS2-BBJ median r_g_=0.885, KCPS2-TWB median r_g_=0.926) and were in general higher than the r_g_ between EAS and EUR (KCPS2-UKB median r_g_=0.815) for these traits (Figure S7, Table S9).

**Figure 4.**
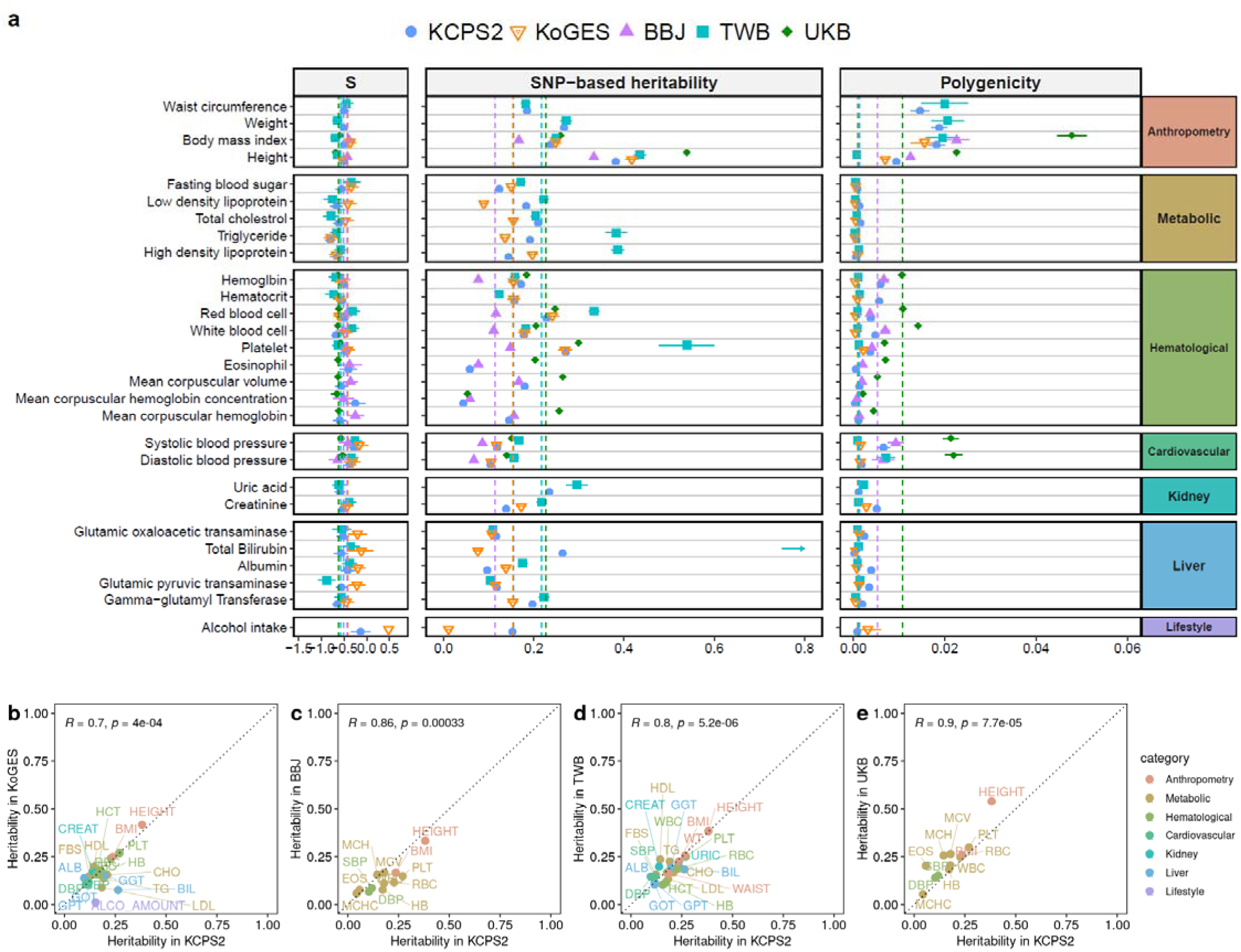
| Genetic architecture of complex traits across KCPS2, BBJ, TWB, and UKB. **(a)**The dots represent posterior means and horizontal bars represent standard errors of the parameters for each trait. The vertical dashed line shows the median of the estimates across traits. Full results are shown in Table S8. **(b-e)** Pearson correlations of SNP-heritability between KCPS2 and KoGES (b), BBJ (c), TWB (d), and UKB (e) across the traits shown in a, except for TWB. For heritability in TWB shown in (d), we used the heritability estimates reported by Chen and colleagues.^15^ The comparisons between KCPS2 heritability estimates and TWB heritability estimates using SbayesS and KCSP2 LD matrix are shown in Table S6b. Data are presented as posterior means of SNP-heritability. The trait categories are indicated by different colors labeled with their trait names. For the 8 traits available in all five studies, we observed high correlations of heritability between KCPS2 and the other biobanks: KoGES (Pearson correlation r=0.99, 95% confidence interval [CI]: 0.97-1.00), BBJ (r=0.93, 95% CI: 0.64-0.99), TWB (r=0.93, 95% CI: 0.65-0.99), and UKB (r=0.97, 95% CI: 0.82-0.99).

### Fine-mapping and colocalization analysis

To identify potential causal variants, we performed a single-population fine-mapping using SuSiE^37^ in KCPS2. Specifically, we fine-mapped 26 traits associated with the region spanning *ALDH2* on chromosome 12 (± 500kb from rs671, which is known to be functionally related to alcohol metabolism). 1,476 variants in this region were fine-mapped to a total of 56 credible sets, among which 17 contain exactly one variant (median 17.5, range 1-470 variants) (Table S10). rs671, a non-synonymous SNP associated with alcohol metabolism and alcohol intake (rs671-A, Beta=-0.59, P=1.9×10^-2658^), had a posterior inclusion probability (PIP) of greater than 90% for 8 traits including alcohol intake, GOT, GPT, GGT, SBP, DBP, coffee intake, and triglyceride (Figure 5a-b). For alcohol intake, we found seven credible sets with exactly one variant, all with PIP=1, including rs671, rs555501971, rs141043717, rs61055528, rs149178839, rs11066008, and rs550463060; in a sensitivity analysis setting the maximum number of credible sets to one (L=1), only rs671 remained in the credible set (Table S11).

**Figure 5.**
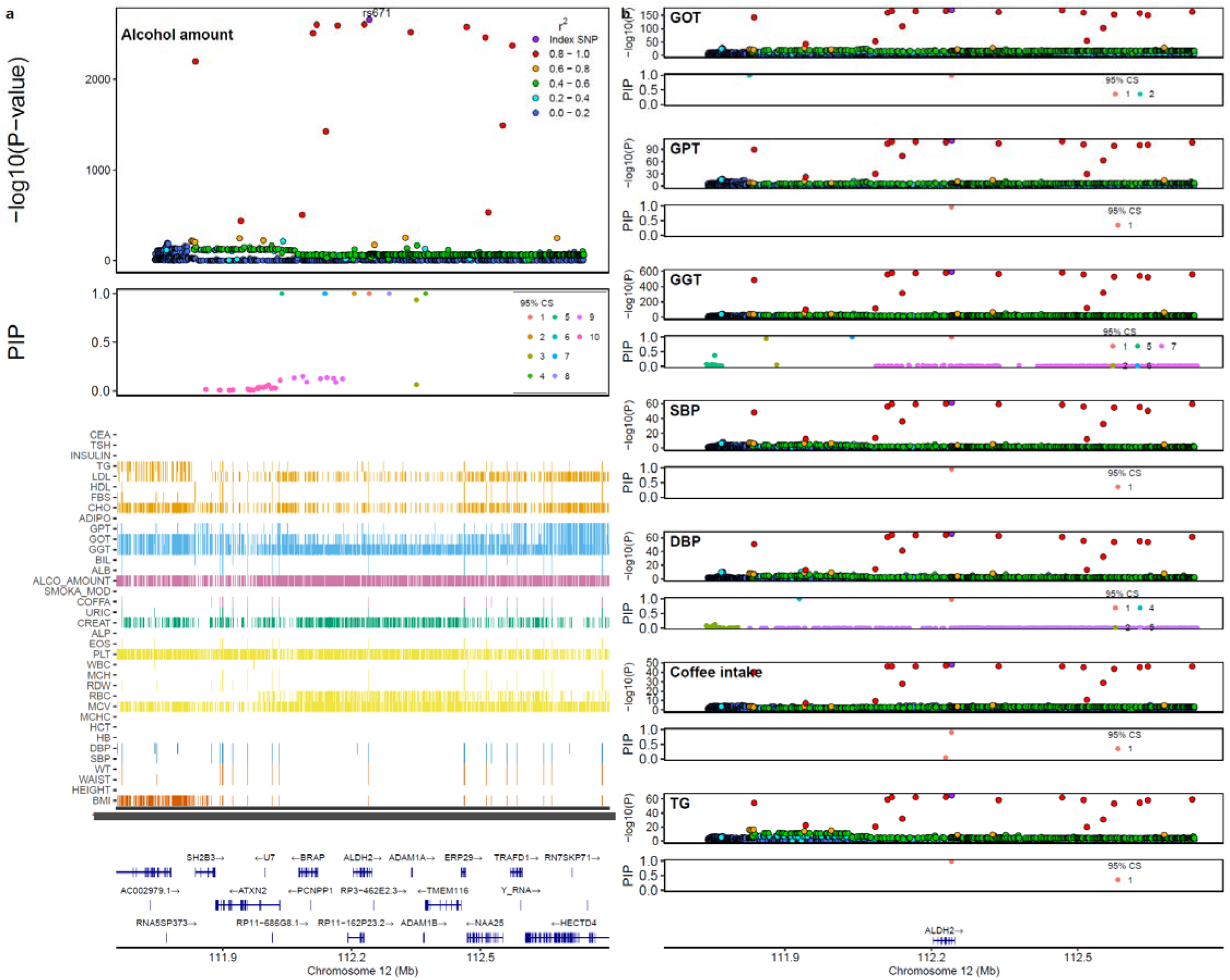
| Fine-mapping and colocalization analysis of ALDH2 region in KCPS2. **(a)** Association between ALDH2 (12q24.12) and alcohol intake in KCPS2. Colors in the Manhattan panels represent r2 values to the lead variant rs671. In the posterior inclusion probability (PIP) panels, only fine-mapped variants in the 95% credible sets (CS) are colored. Heatmap represents significant variants (P<5.0×10^−8^) for the other quantitative traits. **(b)** Colocalization analysis between alcohol intake and seven traits that showed PIP>0.9 for rs671 was done in the same region. Coloc.PP4 represents the posterior probability of colocalization at the specified region. All seven traits shown here had coloc.PP4=1 with alcohol intake. Each regional plot shows associations of each locus for the most significantly associated trait, which was all rs671 with PIP>0.9. Full fine-mapping and colocalization analysis results are shown in Table S10.

To assess the sensitivity of these results to analytic approach, we used conditional and joint analysis (COJO)^38^ to conduct association analyses in the ALDH2 region conditional on rs671 and perform a stepwise selection analysis. These approaches identified independent signals consistent with each other and the original SuSiE analysis (Table S10).

To examine the mechanisms underlying these pleiotropic associations, we performed colocalization by pairing GWAS for alcohol intake with GWAS for each of traits where the PIP of rs671 was greater than 90%. These traits included liver enzymes (GOT, GPT, GGT), blood pressure (SBP, DBP), coffee intake, and triglyceride. rs671 was colocalized between alcohol intake and all of these traits with PP4=100%, supporting a shared causal variant for these two traits at the locus (Figure 5b).

## Discussion

In this study, we identified novel quantitative trait loci for 36 complex traits and investigated the genetic architecture of complex traits in 153,950 Korean individuals. Our analysis discovered 301 novel genetic loci that were not reported in previous GWAS related to the corresponding trait. We also demonstrated widespread pleiotropy and variants near *ALDH2* were associated with 26 traits. Meta-analysis of 21 traits across KCPS2, KoGES, BBJ, TWB, and UKB identified 4,588 loci that were not significant in any of the other four contributing GWAS. We compared the genetic architecture of these traits in KCPS2, KoGES, BBJ, TWB, and UKB, and pinpointed one of the most pleiotropic regions (*ALDH2*) through fine-mapping, which colocalized with high probability with a diverse set of traits such as liver enzyme values.

Our study underscores the importance of enhancing the ancestral diversity and sample size of GWAS samples to facilitate genetic discovery and provide insights into the biological mechanisms of human quantitative complex traits. We discovered 301 novel loci that have lower median MAF in European ancestry individuals than in East Asian populations, which was enabled by leveraging samples from diverse ancestry groups. In particular, the novelty rate was high for TSH (32 out of 100 loci) in KCPS2. The most strongly associated lead variant with TSH, rs10799824-A (Beta=-0.14, P=2.98×10^-139^), was previously reported in GWAS of TSH^39^ (Beta=-0.11, P=4.0×10^-21^) and strict autoimmune hypothyroidism17 (Odds ratio=0.87, P=4.1×10^-18^) in European ancestry individuals. Several lead TSH loci were mapped to nearby genes previously linked to thyroid function, including rs13030651-A (Beta=0.035, P=1.09×10^-13^) in the thyroid adenoma associated gene (*THADA*)^40,41^ and rs2160215 (Beta=0.065, P=1.75×10^-51^) in thyrotropin receptor gene (*TSHR*).^42,43^ Notably, a novel missense variant *CD36* p.Pro90Ser (rs75326924-T; Beta=-0.052, P=9.08×10^-11^) found in our TSH GWAS (AF_KCPS2_=0.068) has low frequency in the gnomAD v4.1.0 East Asian genetic ancestry group (MAF=0.03) but is exceedingly rare outside of that group (MAF<10^-5^) and entirely absent from European genetic ancestry groups.^44^ *CD36* (also known as fatty acid translocase [FAT]) facilitates the transport of fatty acids into cells and participates in triglyceride storage.^45^ A study in hypothyroid rats showed a reduced fatty acid absorption in the liver compared to euthyroid rats^46^ and decreased hepatic FAT expression has been demonstrated in rats with postnatal hypothyroidism.^47^ Moreover, recent studies have revealed that *CD36* contributes to the tumorigenesis and development of multiple cancer types by reprogramming the metabolism of glucose and fatty acid,^48–50^ providing new insights for developing potential therapeutic target and prognostic biomarker in the clinical setting. We also found high novelty rate for GWAS of CEA in KCPS2, which recapitulates several known tumor biomarkers in various cancer types, including a non-coding transcript exon variant (rs149037075-T; Beta=0.179, P=4.8×10^-477^) in *ABO*^51,52^ and a missense variant (rs28362459-C; Beta=0.068, P=4.85×10^-128^) in *FUT3* (also known as Lewis gene),^53–56^ consistent with previous CEA GWAS.^57^ Previous studies show that determinants of the blood A and B antigens and Lewis antigens and of CEA share the same glycoprotein carrier molecules,^58,59^ which might explain the association of CEA concentrations with the *ABO* and the *FUT3* locus. Several variants not previously reported in CEA GWAS were mapped to genes with potential role in cancer such as *C15orf39*^60^ (rs143001709; *P*=1.31×10^-8^), *ST6GAL1*^61^ (rs73187787; *P*=3.36×10^-11^), and *CCDC138*^62^ (rs10179849; P=6.55×10^-19^). Further studies are warranted to investigate the potential functional importance of these associations.

As a global effort to broaden the population diversity of genetic studies in East Asia, the KCPS2 GWAS enhanced our understanding of the genetic basis of complex traits in a Korean population. Our genetic correlations across 36 quantitative traits recapitulated known biology, including negative genetic correlations of HDL, adiponectin, and bilirubin with cardiometabolic risk factors^29,31–33^ and positive genetic correlations of smoking, alcohol intake, and hemoglobin with cardiometabolic traits.^35,63^ Notably, most of the significant genetic correlations were consistent with phenotypic correlations, which underscores the robustness and potential of the genetics-based approaches to understand biological architectures of complex traits. Many of the significant findings were consistent with BBJ,^8^ KoGES,^13^ TWB,^15^ and UKB,^64^ suggesting a similar genetic architecture for these quantitative traits within EAS populations and across EAS and EUR populations as shown by our work and previous findings on within-and cross-ancestry genetic correlation analysis.^6,9,15^

Our findings provide opportunities to investigate the genetic architecture of complex traits within East Asian and across continental populations. While similar negative selection patterns were observed across traits and populations, the heritability estimates vary within EAS and across EAS and EUR populations, which may be attributed to several factors such as phenotype data collection, biobank design and environmental influences. For instance, compared to a hospital-based cohort such as BBJ, participants of a population-/community-based cohort such as KCPS2, KoGES, TWB, and UKB may have different distributions of disease-related traits due to healthy-volunteer effects.^65^ Hence, the comparison of heritability estimates across biobanks requires careful consideration of technical differences, potential collider bias, and variability in baseline health status among studies.^6^ Moreover, we demonstrated the correlation in heritability between KCPS2 and TWB improved when the heritability of TWB were replaced by the previously reported estimates^15^ that used LDSC and in-sample LD, especially for metabolic and hematological traits. Thus, in addition to the phenotype heterogeneity, heritability may be affected by different heritability estimation methods and LD matrices. Further research is needed to explore the impact of these factors on genetic architecture comparisons. Nevertheless, our study highlights the importance of increasing genetic diversity to understand genetic architecture of diverse populations, which is crucial to achieve equitable delivery of genomic knowledge to global populations.^6,66,67^

The KCPS2 GWAS facilitated pinpointing causal variants through fine-mapping. For example, a missense variant rs671 (AF_EAS_=0.2254 vs. AF_EUR_=2.4×10^-5^ in non-Finnish EUR populations in gnomAD v4.0.0^44^) was identified as the causal variant in the ALDH2 gene for 8 traits including alcohol intake, GGT, GOT, GPT, SBP, DBP, coffee intake, and triglyceride through fine-mapping. ALDH2 gene is the target of drug for alcoholism which irreversibly inactivate catalytic Cys302 in ALDH2 by carbamylation in the substrate site of the enzyme.^68,69^ Furthermore, our colocalization results suggest that alcohol intake is a causal risk factor for liver enzymes (GGT, GOT, GPT), blood pressure, and triglyceride levels at the rs671 region, which is supported by evidence for causation from previous MR studies among East Asian populations.^34,70,71^ An additional pleiotropic effect of rs671 across dietary habits on foods and beverages (coffee, green tea, yogurt, natto, tofu and fish) was previously reported, with the same directional effects of rs671 on coffee consumption (rs671-A, Beta=0.061, P=3.4e-49) in our study.^72^ Our findings demonstrated the potential of diversifying EAS GWAS to uncover genetic associations that are common in EAS populations but rare in EUR populations, which could not be discovered even with very large European sample sizes. Furthermore, discovery of such variants may help identify targets for prevention and treatment, thus offering equitable access to precision medicine to diverse populations.

We note several limitations to our study. First, we only conducted GWAS of continuous traits due to limited power for disease phenotypes. Further investigation into disease outcomes should be conducted. Second, we conducted fine-mapping in KCPS2 only for a particular locus, which might cause a concern about potential LD tagging effects for observed pleiotropy. Recent studies suggest that multi-ancestry fine-mapping can improve refinement of causal variants by leveraging different LD patterns across ancestries.^73,74^ We will explore these potential extensions in the near future. Third, for the estimation of genetic architecture parameters, in-sample LD was used for KCPS2, BBJ, and UKB but not for TWB. Since we were unable to find publicly available data to estimate LD in a Taiwanese population, we estimated the genetic architecture parameters using the LD matrix based on the 50K individuals from KCPS2. Such disagreement between the genetic associations and the correlation matrix may induce spurious results due to different LD patterns between the Taiwanese population and Korean individuals from KCPS2, even though both are East Asian populations.

Our findings highlight how broadening the population diversity of GWAS samples can aid discovery and post-GWAS analyses. Our results also provide insights into the genetic architecture of complex traits in East Asian populations. By increasing the sample size and ancestral diversity of GWAS samples, our analysis may help identify novel population-specific targets for prevention and treatment, thus offering equitable access to precision medicine to diverse populations.

## Methods

### Study population

The Korean Cancer Prevention Study-II Biobank (KCPS2) is a prospective cohort study based in Korea with genotype data and measurements of a wide range of phenotypes collected from 153,950 subjects (Figure 1). Participants in KCPS2 undertook routine health assessments at nationwide health promotion centers between 2004 and 2013. Approximately 90% of participants were recruited from the Seoul and Gyeonggi regions, where about 40% of the South Korean population resides (around 19 million people). Mean age of participants at recruitment was 41.7 yr old, and 40% were female. The study design and recruitment have been described in detail previously.^22^ KCPS2 collects extensive phenotypes including demographics, socioeconomic status, environmental exposures, lifestyle, dietary habits, family history and self-reported disease status through structured questionnaires. The anthropometric measures as well as blood and urine samples were collected at recruitment, and several biomarkers were assayed subsequently. All participants in the KCPS2 were genotyped using either the Illumina Global Screening Array (GSA) v2.0 (78,260 samples) or the Korean Chip array v1 (90,245 samples). All participants provided written informed consent before participation.

Quality control (QC) and imputation were conducted separately for each of the two SNP arrays. First, SNPs with low call rate (<95%) were filtered out, along with samples with low call rate (<98%), gender discrepancy, excessive heterozygosity, excessive singletons, and duplicates. Additionally, SNPs with Hardy-Weinberg equilibrium p-value <10-4 or minor allele frequencies (MAF) <0.01 were excluded. Following QC, the data were phased using SHAPEIT4^23^ and imputed using IMPUTE5^24^ with 1000 Genomes Project Phase 3 data. Variants with imputation INFO <0.8 were excluded after imputation. The two imputed data of GSA chip and the Korean chip were then merged, resulting in a total of 6,809,738 overlapping variants.

### Korean Genome and Epidemiology Study (KoGES)

We used KoGES GWAS summary statistics from Nam et al. (https://zenodo.org/record/7042518).^13^ KoGES is a population-based prospective cohort study comprising 72,000 Korean men and women who are recruited from the national health examinee registry at baseline. Mean age of participants at recruitment was 54.15 yr old, and 64% were female. Genome-wide association tests were performed for 76 phenotypes, using linear or proportional odds logistic mixed models implemented in SAIGE (v0.44.5)^25^ or POLMM^75^ adjusted for top 10 principal components (PC), age, sex, and adjustment of assessment details such as cohort and year of examination. We extracted 22 quantitative traits from KoGES that matched the quantitative traits we analyzed in KCPS2: albumin (ALB), alcohol amount (ALCO_AMOUNT), gamma-glutamyl Transferase (GGT), glutamic oxaloacetic transaminase (GOT), body mass index (BMI), creatinine (CREAT), diastolic blood pressure (DBP), glutamic pyruvic transaminase, (GPT), fasting blood sugar (FBS), hemoglobin (HB), high density lipoprotein (HDL), height (HEIGHT), hematocrit (HCT), low density lipoprotein (LDL), platelet (PLT), red blood cell (RBC), systolic blood pressure (SBP), total bilirubin (BIL), total cholesterol (CHO), triglyceride (TG), waist circumference (WC), and white blood cell (WBC). The quantitative traits used in the KoGES GWAS were inverse rank-based normal transformed.

### Biobank Japan (BBJ)

We used GWAS summary statistics from BBJ from Sakaue et al. (https://pheweb.jp/).^9^ BBJ is a hospital-based cohort, consisting of 179,000 Japanese participants. The mean age of participants at recruitment was 63.0 years, and 46.3% were female. Genome-wide association tests were conducted for 220 phenotypes, using linear or logistic mixed models implemented in SAIGE (v0.37)^25^ or BOLT-LMM (v.2.3.4).^76^ The measured values of each quantitative trait were adjusted for age, age^2^, sex, age by sex interaction, age^2^ by sex interaction, and the top 20 PCs, and the residuals were then transformed by rank-based inverse normalization. We extracted 27 quantitative traits from BBJ that matched the quantitative traits we analyzed in KCPS2: LDL, TG, HDL, CHO, FBS, GOT, GPT, GGT, BIL, ALB, CREAT, uric acid (URIC), alkaline phosphatase (ALP), HB, HCT, mean corpuscular hemoglobin (MCH), mean corpuscular hemoglobin concentration (MCHC), mean corpuscular volume (MCV), RBC, WBC, PLT, eosinophil (EOS), SBP, DBP, weight (WT), HEIGHT, and BMI.

### Taiwan Biobank (TWB)

TWB is a population-based prospective cohort study of the Taiwanese population, comprising 102,900 participants. The mean age of participants at recruitment was 50.0 years, and 64% were female. We used two sets of TWB GWAS summary statistics from Chen et al.^15^ for the meta-analysis and the genetic architecture analysis, respectively. For the meta-analysis, we used the TWB GWAS summary statistics, using linear mixed models implemented in Regenie (v1.0.5.4)^77^ adjusted for age, age^2^, sex, age by sex interaction, age^2^ by sex interaction, and top 20 PCs. We extracted 23 quantitative traits from BBJ that matched the quantitative traits we analyzed in KCPS2: LDL, TG, HDL, CHO, FBS, GOT, GPT, GGT, BIL, ALB, CREAT, URIC, HB, HCT, RBC, WBC, PLT, SBP, DBP, WT, HEIGHT, BMI, and WAIST. All phenotypes used in the TWB GWAS were inverse rank-based normal transformed. For the genetic architecture analysis, we used TWB GWAS summary statistics using linear regression implemented in PLINK2,^78^ as the previously reported genetic architecture estimates we compared against were also based on linear association test statistics.

### UK Biobank (UKB)

The UKB project is a population-based prospective cohort that recruited approximately 500,000 participants from across the United Kingdom. The mean age of participants at recruitment was 56.8 years, with 53.8% identifying as female. All GWAS summary statistics from UKB used in this study were publicly available at: https://pan.ukbb.broadinstitute.org/, generated and released by Karczewski et al.^16^ For our analysis, we used the EUR ancestry estimates from the Pan-UKB data. We included 29 GWAS from UKBB: Height, BMI, HB, HCT, RBC, WBC, PLT, SBP, DBP, WT, LDL, TG, HDL, CHO, FBS, GGT, BIL, ALB, CREAT, ALCO_AMOUNT, GOT, GPT, URIC, ALP, MCH, MCHC, MCV, EOS, and WAIST. All phenotypes used in these GWAS were inverse rank-based normal transformed.

### Genome-wide association analysis in KCPS2

We performed GWAS on 36 quantitative traits including anthropometric measures and biomarkers spanning 8 categories (metabolic, liver, thyroid hormone, tumor marker, kidney, hematological, cardiovascular, arthrometry, and lifestyle factors). For each trait, we excluded samples with measurements that were more than 6 standard deviations away from the sample average.

We used linear mixed models implemented in SAIGE (v.1.1.9)^25^ for association testing, controlling for age, sex, 10 PCs, and SNP array. The SAIGE method contains two main steps: in step 1, we used a subset of linkage disequilibrium (LD)-pruned variants with R^2^<0.2 (158,729 variants) to obtain the genetic relationship matrix.

We included age, sex, 10 PCs, and SNP array as covariates in step 1. Single-variant association testing was performed in step 2 where the phenotypes were inverse rank-based normal transformed and leave-one-chromosome-out scheme to remove the proximal contamination. We used FUMA^28^ with 1000 Genome Project Phase 3^79^ EAS samples as LD reference to identify independent genome-wide significant loci (p<5×10^-8^) for each trait, window size of 500 kb, and LD threshold R^2^ of 0.1.

Linkage disequilibrium score regression (LDSC)^80^ was applied to estimate cross-trait genetic correlations (rg) in KCPS2. We ran stratified-LDSC (S-LDSC)^81^ with a full baseline-LD v1.2 model^81^ to compute LDSC intercept. To correctly specify effective sample size in LDSC or S-LDSC analysis, we used GWAS summary statistics generated from simple linear regression models instead of linear mixed models, which have a different effective GWAS sample size than the study sample size.^82^ We ran linear regression using PLINK2^78^ for association testing, controlling for age, sex, 10 PCs, and SNP array in unrelated KCPS2 samples. Specifically, we removed second degree or more closely related individuals using the software KING.^83^ All phenotypes used in these GWAS were inverse rank-based normal transformed.

### Novel association identification

We mapped each trait to a term in the Experimental Factor Ontology (EFO) each trait (Table S3). For each of the independent loci we identified to be associated with a given trait, we queried the Open Target Genetics database (release 22.09)^27,84^ for each of the independent loci we identified to be associated with a given trait to identify any previously reported associations (with the same EFO term or category, see below). We considered a trait-associated locus as novel when the locus with ±500 kb flanking window did not include any lead variants that were previously reported with the same EFO term. Given the widespread pleiotropy and phenotypic heterogeneity,^85^ we may overcount novel associations. We therefore also used EFO categories, which are more generic than EFO terms to evaluate novelty. For example, “body height” (EFO_0004339) is an EFO term that maps to the broader EFO category of “body measurement” (EFO_0004324) by GWAS Catalog.^26^ We further exhaustively searched for previous reports of genetic association in a given trait using the GWAS Catalog, which might not be included in the Open Target Genetics database. Since the recent GWAS results of height^86^ and TSH^87,88^ were not listed in the GWAS Catalog at the time of the curation, we additionally excluded variants that were genome-wide significant in the GWAS. Since the results by KoGES^13^ and TWB^15^ were not listed in the Open Target Genetics database or the GWAS Catalog at the time of evaluation for the novel association, we additionally excluded loci with ±500 kb flanking window that include any previous associations reported by KoGES or TWB. A flow chart illustrating the process for identifying novel loci is demonstrated in Supplementary Figure 8.

### Evaluation of gene pleiotropy

We investigated gene pleiotropy, where a gene affects multiple traits in KCPS2. We defined the degree of pleiotropy as the number of significant associations per gene (p<5×10^-8^). The list of genes mapped to each SNP in KCPS2 GWAS results was taken from FUMA^28^ to map SNPs in GWAS results to a gene with the 1000 Genome Phase 3^79^ EAS reference panel. We then quantified the degree of pleiotropy per gene by aggregating and counting the number of genome-wide significant associations across 36 traits.

### Meta-analysis of EAS and EUR GWAS

We conducted meta-analysis of 21 traits across KCPS2, KoGES, BBJ, TWB, and UKB (European ancestry samples) to further identify novel loci across East Asian and European ancestry populations. We implemented inverse-variance-weighted fixed-effect meta-analysis using METAL.^89^ We retained variants presented in at least one of the studies in this meta-analysis for loci discovery in the EAS and EUR populations. We then used FUMA^28^ to identify genome-wide significant loci in the meta-analysis after clumping variants with p-values<5×10^-8^, window size of 5 Mb, and LD threshold R^2^ of 0.1. We identified the association as novel if none of the variants within the locus reached genome-wide significance (P< 5×10^-8^) in KoGES, BBJ, TWB, or UKB GWAS.

### Genetic architecture of complex traits in KCPS2, KoGES, BBJ, TWB, and UKB

We used SbayesS^90^ to estimate the SNP-based heritability (lil^2^), polygenicity (π; proportion of SNPs with nonzero effects), and the relationship between minor allele frequency (MAF) and SNP effects (S parameter) for 36 traits in KCPS2. We constructed a full LD correlation matrix based on 50K unrelated individuals from KCPS2 and shrunk the matrix to ignore small LD correlations due to sampling variance using the shrinkage method from Wen and Stephens (2010).^91^ To calculate the LD matrix shrinkage estimate, we used a genetic map for East Asian populations, with the effective population sample size of 12,239,^79^ while using the default shrinkage cutoff (10^-5^). We then compared the genetic architecture of KCPS2 with KoGES, BBJ, TWB, and UKB across six categories including anthropometry, cardiovascular, hematological, kidney, liver, and metabolic traits (among which 22 traits overlap between KCPS2 and KoGES, 12 traits overlap between KCPS2 and BBJ/UKB, 23 traits overlap between KCPS2 and TWB, and 8 traits available in all biobanks: height, body mass index, platelet, red blood cell, white blood cell, hemoglobin, systolic blood pressure, and diastolic blood pressure). When comparing the median SNP-heritability of trait categories between two studies, we restricted the comparison to the same list of traits within each category for that specific biobank pair and performed a Wilcoxon signed-rank test to assess statistical significance. For BBJ and UKB, we used the previously reported genetic architecture parameter estimates,^66^ which were constructed using GWAS summary statistics generated from linear regression models and in-sample LD for the corresponding unrelated population. For a fair comparison of these parameters between KCPS2, BBJ, TWB, and UKB, we applied SbayesS to GWAS summary statistics generated from linear regression models in unrelated KCPS2 and TWB, instead of linear mixed models. Since summary statistics from linear regression models were not publicly available for KoGES, we used linear mixed models instead. For TWB, we estimated the genetic architecture parameters using the LD matrix based on the 50K individuals from KCPS2 because we were unable to find publicly available data to estimate LD in a Taiwanese population. As we noted in our limitation, presumably because unmatched LD was used for TWB, we found overestimation of SbayesS heritability estimation in TWB. This overestimation was magnified in total bilirubin on chromosome 2 (lil^2^ =1.12), which is a Mendelian locus, harboring the *UGT1A1* gene. Thus, we removed chromosome 2 when estimating correlations between heritability in KCPS2 and TWB (Supplementary Figure 6b).

### Cross-biobank genetic correlation

To estimate cross-biobank genetic effect correlations within EAS (KCPS2-KoGES, KCPS2-BBJ, and KCPS2-TWB), we used LDSC^80^ to estimate r_g_ using the 1000 Genomes phase 3 EAS reference panel. We used Popcorn (v.1.0)^92^ to estimate cross-biobank genetic-effect correlation between KCPS2 and UKB GWAS with precomputed cross-population scores for EUR and EAS 1000 Genomes Project populations provided by the authors. For a fair comparison, we restricted to HapMap3 SNPs that were shared across all five biobanks. We applied the analysis to traits with heritability calculated by LDSC or Popcorn >0.01 and their GWAS summary statistics generated from linear mixed models from all biobanks which were publicly available.

### Fine-mapping and colocalization analysis

We fine-mapped one of the most pleiotropic regions identified by GWAS of the 36 traits above, a 500kb region flanking *ALDH2* in KCPS2. We applied SuSiE^37^ to GWAS summary statistics and in-sample LD on 1,476 SNPs in this region. We implemented colocalization analysis to further investigate whether two traits share a causal variant. We applied *coloc.susie*^93^ which allows multiple signals to be distinguished using SuSiE, and then performed colocalization analysis on all possible pairs of signals between the traits. We performed colocalization analysis in a 500kb window centered on an identified causal variant between alcohol intake and the other traits with PIP of rs671 being greater than 0.9 from fine-mapping results. We reported posterior probability of colocalization (PP4) for each of these pairs at the specified region. We applied LocusZoom^94^ to visualize the colocalization analysis.

#### Acknowledgements

We acknowledge the participants of KCPS2 and the management team and leadership of KCPS2 for their outstanding support in collecting samples and clinical data. We thank KoGES, BBJ, TWB, and UKB for providing resources and releasing the GWAS summary statistics, which made this study possible. K.J.J acknowledged support from the Basic Science Research Program through the National Research Foundation of Korea funded by the Ministry of Education (RS-2023-00239122). R.D. was supported by R01 GM148494. A.L.P was supported by R01 HG006399. A.R.M. was supported by funding from the National Institutes of Health (K99/R00MH117229) and funding from a Broad Next Generation Fund. P.K. was supported by U01CA261339.

## Data availability

GWAS summary statistics in this study will be made publicly available prior to publication. Data sources for other publicly available GWAS summary statistics are available in Supplementary Table 13. The summary statistics for KoGES used in this study were downloaded from the KoGES Zenodo (https://zenodo.org/record/7042518), BBJ summary statistics from the Biobank Japan PheWeb (https://pheweb.jp/), TWB summary statistics from GWAS Catalog (https://www.ebi.ac.uk/gwas/publications/38116116), and summary statistics for Europeans in UKB were downloaded from Pan-UK Biobank (https://pan.ukbb.broadinstitute.org/).

## Code availability

We used publicly available software for the analyses. The software used is listed and described in the Methods section of our manuscript. Analysis code used in this manuscript is publicly available at https://github.com/yonhojee/KCPS2-GWAS.

## Supplementary Figures

**Supplementary Figure 1.**
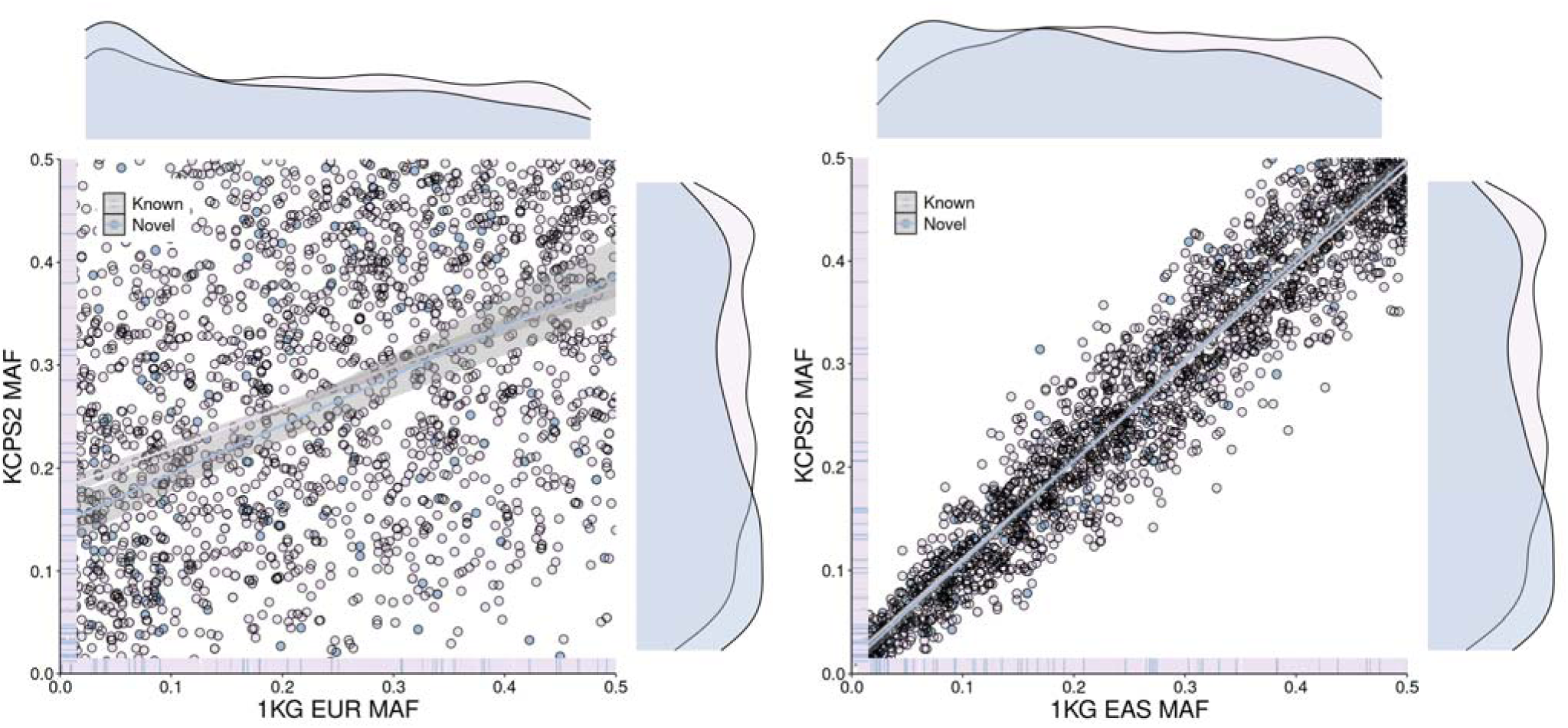
MAF comparisons of 2,962 independent genome-wide significant loci in KCPS2 with MAF in 1000 Genomes Project (1KG) European (EUR) and East Asian (EAS) population. The median MAF of the novel loci was 0.118 in 1KG EUR (paired t test P=2.2e-16) and 0.202 (paired t-test P=3.8e-4) in 1KG EAS.

**Supplementary Figure 2.**
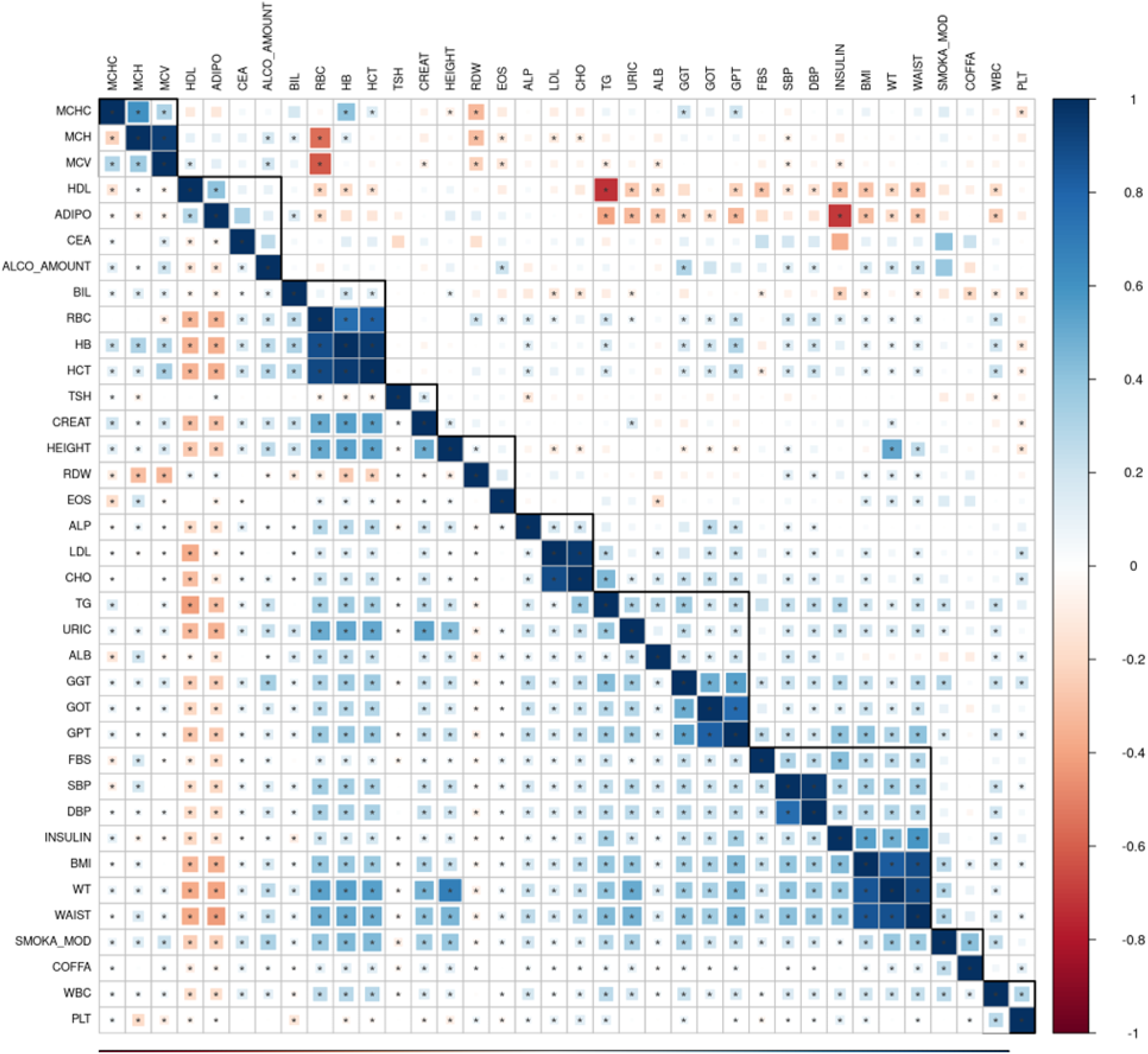
Pairwise genetic correlations (r_g_, upper diagonal) and phenotypic correlations (r_p_, lower diagonal) between the 36 traits in KCPS2. r_g_ was estimated using bivariate LDSC based on association test statistics from linear regression. Significant r_g_ and r_p_ after false discovery rate (FDR<0.05) correction is indicated by an asterisk sign (two-sided Wald test). The complete set of r_g_ and r_p_, is available in Table S5.

**Supplementary Figure 3.**
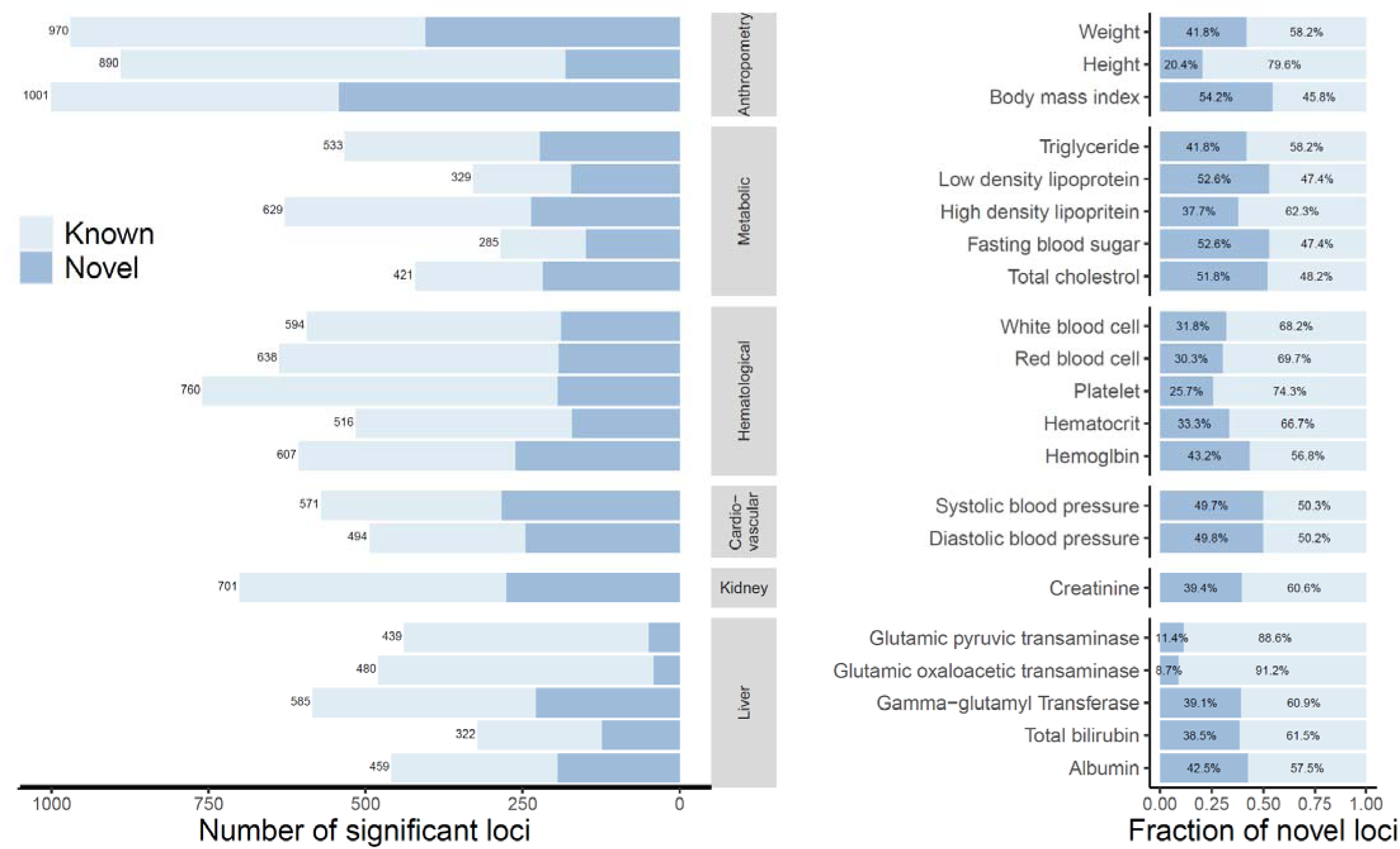
Number of known and novel variants identified in the meta-analysis across Korean Cancer Prevention study-II (KCPS2), Biobank Japan (BBJ), Korean Genome and Epidemiology Study (KoGES), Taiwan Biobank (TWB), and UK Biobank (UKB). We identified the association as novel if none of the variants within the locus reached genome-wide significance (P< 5×10^-8^) in KoGES, BBJ, TWB, or UKB GWAS.

**Supplementary Figure 4.**
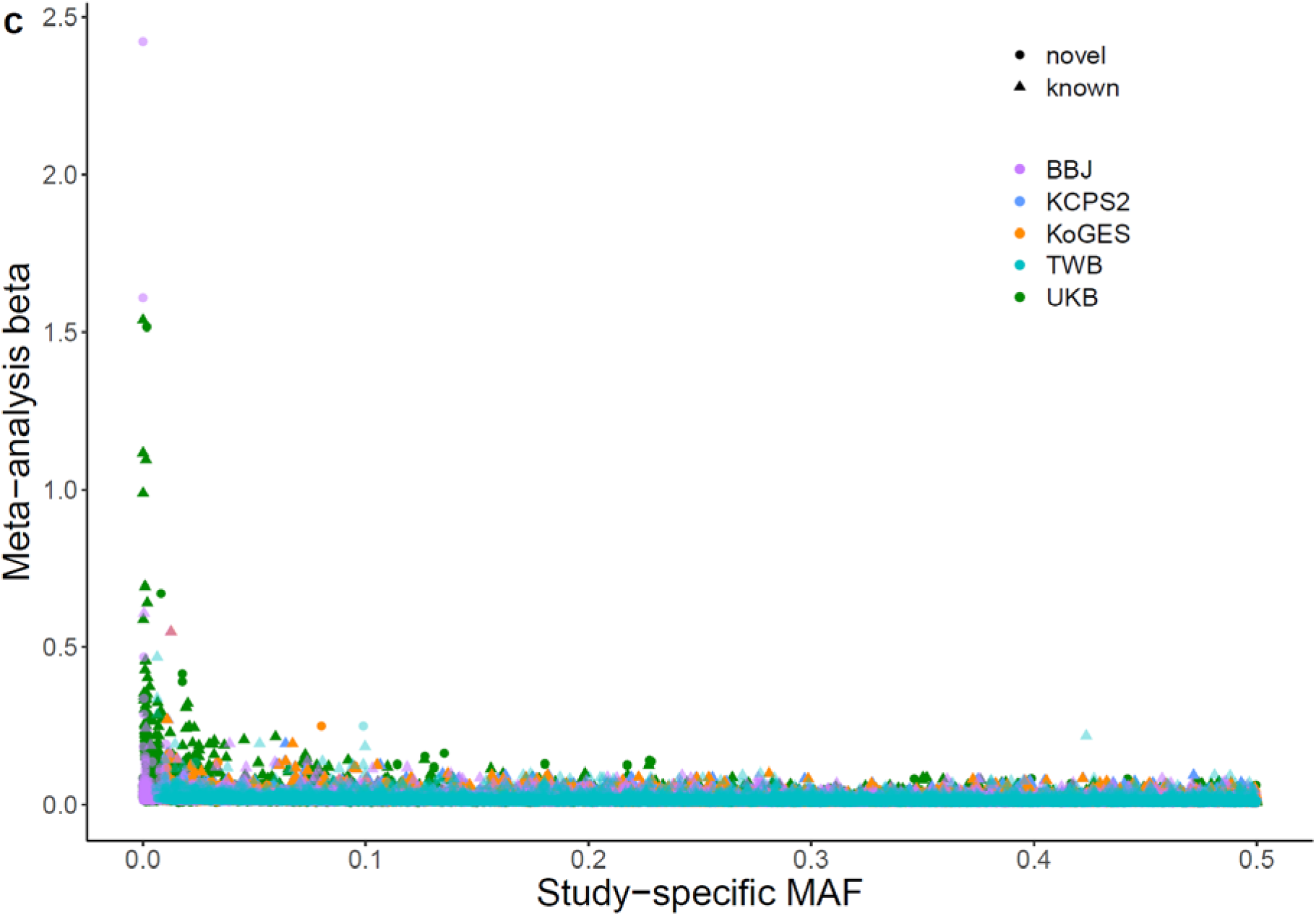
Comparisons of study-specific minor allele frequencies and effect sizes estimated from the multi-ancestry meta-analysis. We identified the association as novel if none of the variants within the locus reached genome-wide significance (*P*< 5×10^-8^) in KoGES, BBJ, TWB, or UKB GWAS.

**Supplementary Figure 5.**
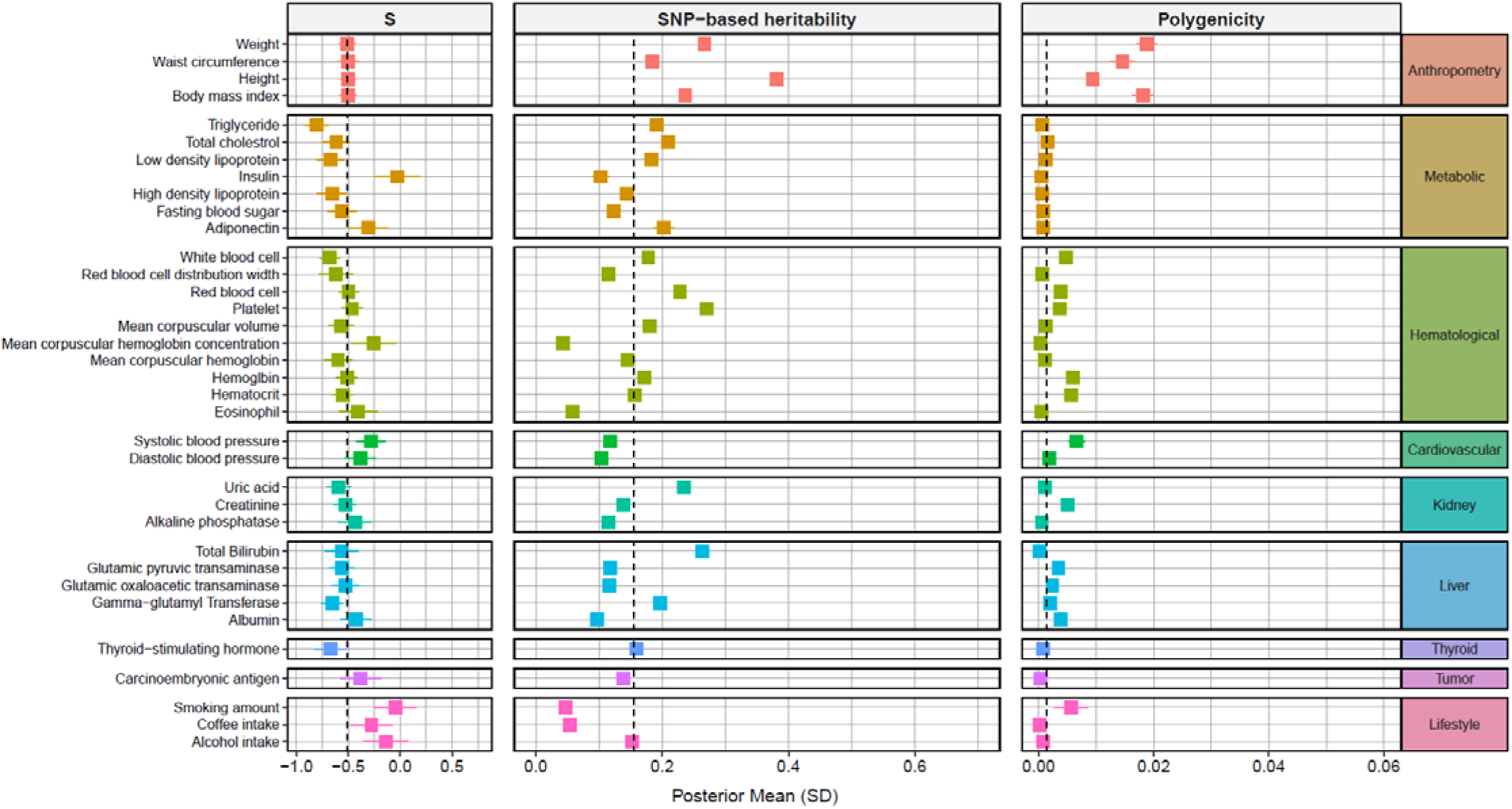
Genetic architecture of 36 traits in Korean Cancer Prevention study-II (KCPS2). The dots represent posterior means and horizontal bars represent standard errors of the parameters for each trait. The vertical dashed line shows the median of the estimates across traits.

**Supplementary Figure 6.**
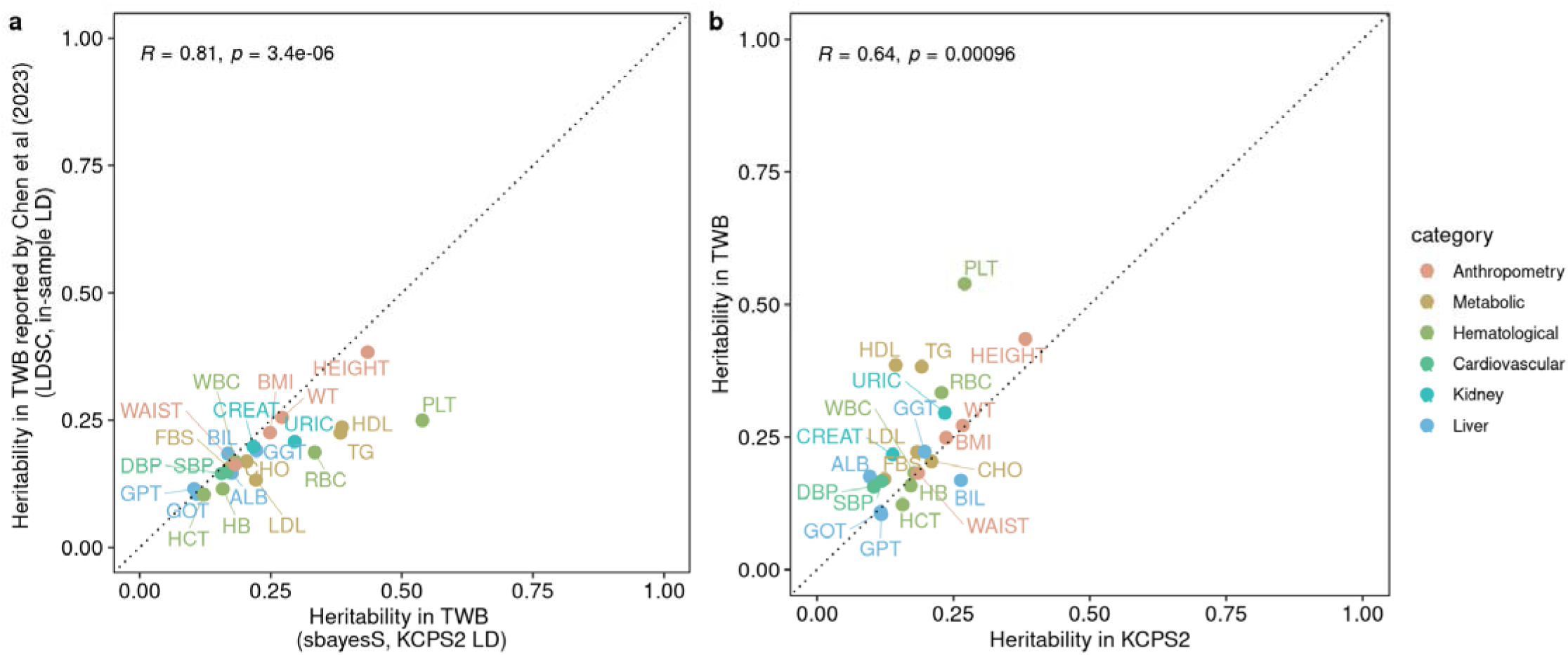
Comparisons of KCPS2 heritability estimates with TWB heritability estimates using different methods and LD reference panel. A) Comparisons between TWB heritability estimates using SbayesS and KCSP2 LD matrix (X-axis) and TWB heritability estimates reported by Chen et al., (2023)^15^ using LDSC and in-sample LD from TWB (Y-axis). B) Comparisons between KCPS2 heritability estimates (X-axis) and TWB heritability estimates using SbayesS and KCSP2 LD matrix (Y-axis). For panel (b), we removed chromosome 2 heritability estimate for total bilirubin in TWB, which led to heritability being greater than 1 (*h^2^_g_*_g_=1.12) due to the Mendlian locus in chromosome 2. If we did not remove chromosome 2 heritability estimate, the correlation between heritability in KCPS2 and TWB was 0.56 (P=0.0056).

**Supplementary Figure 7.**
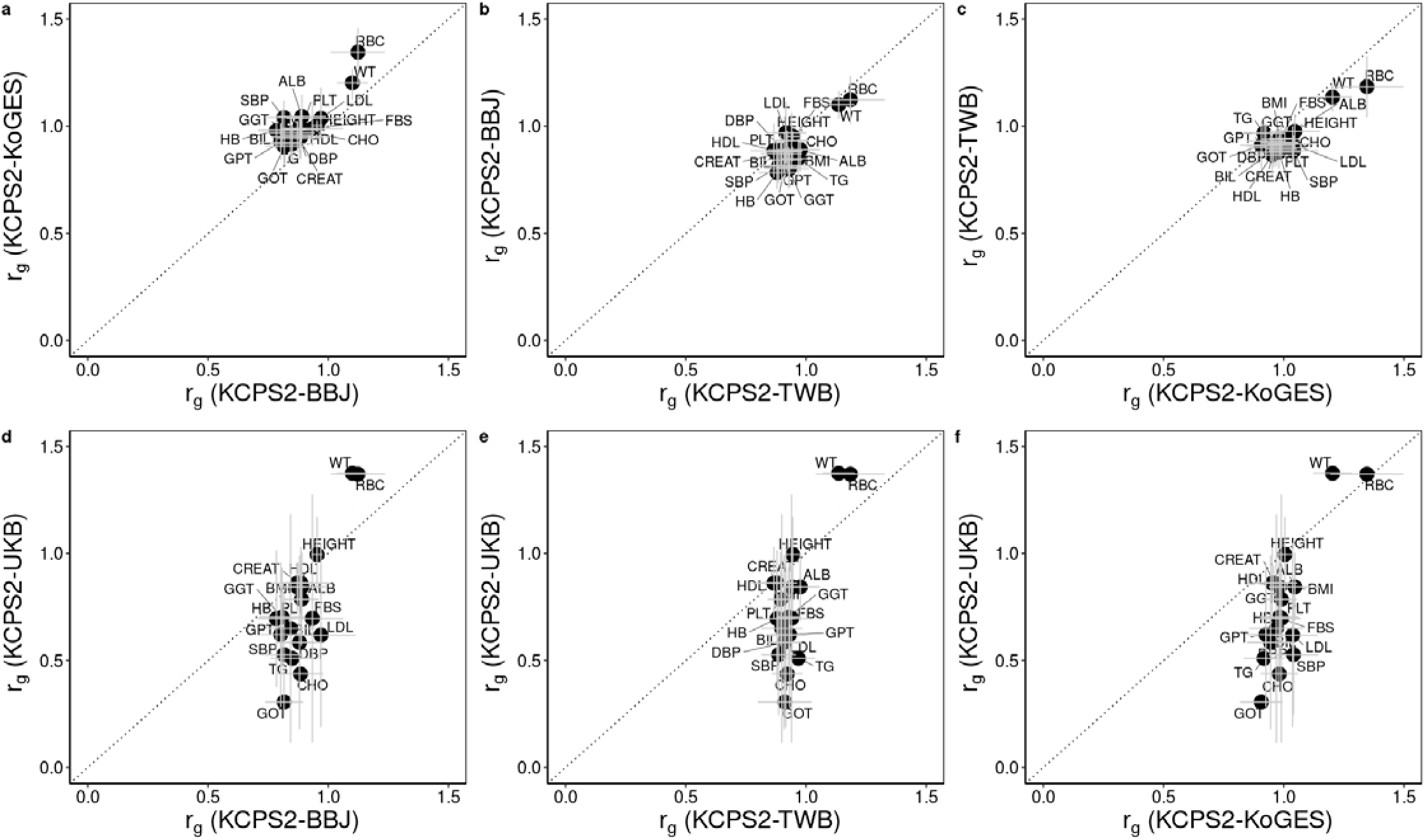
Comparison of within- and cross-biobank genetic correlation estimates for 21 quantitative traits in KCPS2, KoGES, BBJ, TWB, and UKBB (r_g_). (a-c) The r_g_ estimates within EAS were computed in LDSC^80^ using 1000 Genomes Project EAS reference panel. (d-f) The cross-biobank population genetic effect correlations between KCPS2 and UKB were estimated in Popcorn (v.1.0)^92^ using precomputed cross-population scores for EUR and EAS 1000 Genomes Project populations (Tables S9).

**Supplementary Figure 8.**
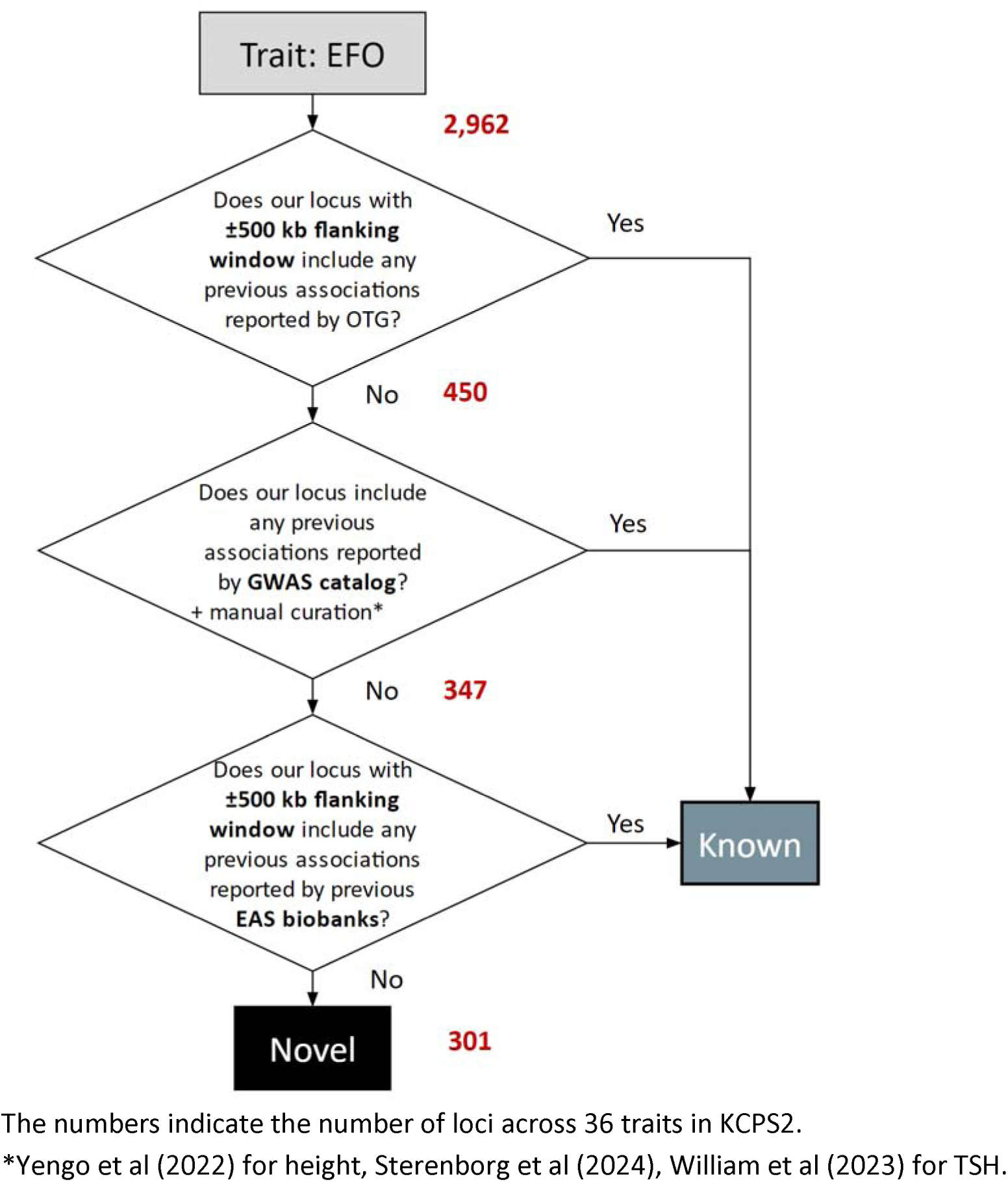
Flow chart of identifying novel loci in KCPS2.

## References

1. Tam V, Patel N, Turcotte M, Bossé Y, Paré G, Meyre D. Benefits and limitations of genome-wide association studies. Nat Rev Genet. 2019 Aug;20(8):467–484.

2. Abdellaoui A, Yengo L, Verweij KJH, Visscher PM. 15 years of GWAS discovery: Realizing the promise. Am J Hum Genet. 2023 Feb 2;110(2):179–194.

3. Chatterjee N, Shi J, García-Closas M. Developing and evaluating polygenic risk prediction models for stratified disease prevention. Nat Rev Genet. Nature Publishing Group; 2016 Jul;17(7):392–406.

4. Lambert SA, Abraham G, Inouye M. Towards clinical utility of polygenic risk scores. Hum Mol Genet. 2019 Nov 21;28(R2):R133–R142.

5. Fatumo S, Chikowore T, Choudhury A, Ayub M, Martin AR, Kuchenbaecker K. A roadmap to increase diversity in genomic studies. Nat Med. Nature Publishing Group; 2022 Feb;28(2):243–250.

6. Martin AR, Kanai M, Kamatani Y, Okada Y, Neale BM, Daly MJ. Clinical use of current polygenic risk scores may exacerbate health disparities. Nat Genet. Nature Publishing Group; 2019 Apr;51(4):584–591.

7. Akiyama M, Okada Y, Kanai M, et al. Genome-wide association study identifies 112 new loci for body mass index in the Japanese population. Nat Genet. Nature Publishing Group; 2017 Oct;49(10):1458–1467.

8 . Kanai M, Akiyama M, Takahashi A, et al. Genetic analysis of quantitative traits in the Japanese population links cell types to complex human diseases. Nat Genet. Nature Publishing Group; 2018 Mar;50(3):390–400.

9. Sakaue S, Kanai M, Tanigawa Y, et al. A cross-population atlas of genetic associations for 220 human phenotypes. Nat Genet. Nature Publishing Group; 2021 Oct;53(10):1415–1424.

10. Kim T, Park AY, Baek Y, Cha S. Genome-Wide Association Study Reveals Four Loci for Lipid Ratios in the Korean Population and the Constitutional Subgroup. PLOS ONE. Public Library of Science; 2017 Jan 3;12(1):e0168137.

11. Cho H-W, Jin H-S, Eom Y-B. A Genome-Wide Association Study of Novel Genetic Variants Associated With Anthropometric Traits in Koreans. Frontiers in Genetics [Internet]. 2021 [cited 2024 Jan 26];12. Available from: https://www.frontiersin.org/articles/10.3389/fgene.2021.669215

12. Park J sung, Kim Y, Kang J. Genome-wide meta-analysis revealed several genetic loci associated with serum uric acid levels in Korean population: an analysis of Korea Biobank data. J Hum Genet. Nature Publishing Group; 2022 Apr;67(4):231–237.

13. Nam K, Kim J, Lee S. Genome-wide study on 72,298 individuals in Korean biobank data for 76 traits. Cell Genomics. 2022 Oct 12;2(10):100189.

14. Walters RG, Millwood IY, Lin K, et al. Genotyping and population characteristics of the China Kadoorie Biobank. Cell Genom. 2023 Aug 9;3(8):100361.

15. Chen C-Y, Chen T-T, Feng Y-CA, et al. Analysis across Taiwan Biobank, Biobank Japan, and UK Biobank identifies hundreds of novel loci for 36 quantitative traits. Cell Genomics. 2023 Dec 13;3(12):100436.

16. Karczewski KJ, Gupta R, Kanai M, et al. Pan-UK Biobank GWAS improves discovery, analysis of genetic architecture, and resolution into ancestry-enriched effects [Internet]. medRxiv; 2024 [cited 2024 May 2]. p. 2024.03.13.24303864. Available from: https://www.medrxiv.org/content/10.1101/2024.03.13.24303864v1

17. Kurki MI, Karjalainen J, Palta P, et al. FinnGen provides genetic insights from a well-phenotyped isolated population. Nature. Nature Publishing Group; 2023 Jan;613(7944):508–518.

18. Brumpton BM, Graham S, Surakka I, et al. The HUNT study: A population-based cohort for genetic research. Cell Genom. 2022 Oct 12;2(10):100193.

19. Gudbjartsson DF, Helgason H, Gudjonsson SA, et al. Large-scale whole-genome sequencing of the Icelandic population. Nat Genet. 2015 May;47(5):435–444.

20. Bycroft C, Freeman C, Petkova D, et al. The UK Biobank resource with deep phenotyping and genomic data. Nature. Nature Publishing Group; 2018 Oct;562(7726):203–209.

21. Mills MC, Rahal C. The GWAS Diversity Monitor tracks diversity by disease in real time. Nat Genet. Nature Publishing Group; 2020 Mar;52(3):242–243.

22. Jee YH, Emberson J, Jung KJ, et al. Cohort Profile: The Korean Cancer Prevention Study-II (KCPS-II) Biobank. International Journal of Epidemiology. 2018 Apr 1;47(2):385–386f.

23. Delaneau O, Zagury J-F, Robinson MR, Marchini JL, Dermitzakis ET. Accurate, scalable and integrative haplotype estimation. Nat Commun. 2019 Nov 28;10(1):5436.

24. Rubinacci S, Delaneau O, Marchini J. Genotype imputation using the Positional Burrows Wheeler Transform. PLoS Genet. 2020 Nov;16(11):e1009049.

25. Zhou W, Nielsen JB, Fritsche LG, et al. Efficiently controlling for case-control imbalance and sample relatedness in large-scale genetic association studies. Nat Genet. 2018 Sep;50(9):1335–1341.

26. Sollis E, Mosaku A, Abid A, et al. The NHGRI-EBI GWAS Catalog: knowledgebase and deposition resource. Nucleic Acids Res. 2023 Jan 6;51(D1):D977–D985.

27. Ghoussaini M, Mountjoy E, Carmona M, et al. Open Targets Genetics: systematic identification of trait-associated genes using large-scale genetics and functional genomics. Nucleic Acids Res. 2021 Jan 8;49(D1):D1311–D1320.

28. Watanabe K, Taskesen E, Bochoven A van, Posthuma D. Functional mapping and annotation of genetic associations with FUMA. Nat Commun. Nature Publishing Group; 2017 Nov 28;8(1):1826.

29. Perswani P, Ismail SM, Mumtaz H, et al. Rethinking HDL-C: An In-Depth Narrative Review of Its Role in Cardiovascular Health. Current Problems in Cardiology. 2024 Feb 1;49(2):102152.

30. Kawano J, Arora R. The role of adiponectin in obesity, diabetes, and cardiovascular disease. J Cardiometab Syndr. 2009;4(1):44–49.

31. Tang Y-H, Wang Y-H, Chen C-C, Chan C-J, Tsai F-J, Chen S-Y. Genetic and Functional Effects of Adiponectin in Type 2 Diabetes Mellitus Development. International Journal of Molecular Sciences. Multidisciplinary Digital Publishing Institute; 2022 Jan;23(21):13544.

32. Wen G, Yao L, Hao Y, Wang J, Liu J. Bilirubin ameliorates murine atherosclerosis through inhibiting cholesterol synthesis and reshaping the immune system. Journal of Translational Medicine. 2022 Jan 3;20(1):1.

33. Hinds TD, Stec DE. Bilirubin, a Cardiometabolic Signaling Molecule. Hypertension. American Heart Association; 2018 Oct;72(4):788–795.

34. Xu L, Jiang CQ, Cheng KK, et al. Alcohol Use and Gamma-Glutamyltransferase Using a Mendelian Randomization Design in the Guangzhou Biobank Cohort Study. PLoS One. 2015;10(9):e0137790.

35. Auvinen J, Tapio J, Karhunen V, et al. Systematic evaluation of the association between hemoglobin levels and metabolic profile implicates beneficial effects of hypoxia. Sci Adv. 2021 Jul 14;7(29):eabi4822.

36. Zeng J, Vlaming R de, Wu Y, et al. Signatures of negative selection in the genetic architecture of human complex traits. Nat Genet. Nature Publishing Group; 2018 May;50(5):746–753.

37. Wang G, Sarkar A, Carbonetto P, Stephens M. A Simple New Approach to Variable Selection in Regression, with Application to Genetic Fine Mapping. Journal of the Royal Statistical Society Series B: Statistical Methodology. 2020 Dec 1;82(5):1273–1300.

38. 38. Yang J, Ferreira T, Morris AP, et al. Conditional and joint multiple-SNP analysis of GWAS summary statistics identifies additional variants influencing complex traits. Nat Genet. Nature Publishing Group; 2012 Apr;44(4):369–375.

39. Porcu E, Medici M, Pistis G, et al. A Meta-Analysis of Thyroid-Related Traits Reveals Novel Loci and Gender-Specific Differences in the Regulation of Thyroid Function. PLOS Genetics. Public Library of Science; 2013 7;9(2):e1003266.

40. Cancer Genome Atlas Research Network. Integrated genomic characterization of papillary thyroid carcinoma. Cell. 2014 Oct 23;159(3):676–690.

41. Drieschner N, Kerschling S, Soller JT, et al. A domain of the thyroid adenoma associated gene (THADA) conserved in vertebrates becomes destroyed by chromosomal rearrangements observed in thyroid adenomas. Gene. 2007 Nov 15;403(1–2):110–117.

42. Calebiro D, Gelmini G, Cordella D, et al. Frequent TSH receptor genetic alterations with variable signaling impairment in a large series of children with nonautoimmune isolated hyperthyrotropinemia. J Clin Endocrinol Metab. 2012 Jan;97(1):E156–160.

43. Abramowicz MJ, Duprez L, Parma J, Vassart G, Heinrichs C. Familial congenital hypothyroidism due to inactivating mutation of the thyrotropin receptor causing profound hypoplasia of the thyroid gland. J Clin Invest. 1997 Jun 15;99(12):3018–3024.

44. Karczewski KJ, Francioli LC, Tiao G, et al. The mutational constraint spectrum quantified from variation in 141,456 humans. Nature. Nature Publishing Group; 2020 May;581(7809):434–443.

45. Febbraio M, Hajjar DP, Silverstein RL. CD36: a class B scavenger receptor involved in angiogenesis, atherosclerosis, inflammation, and lipid metabolism. J Clin Invest. 2001 Sep;108(6):785–791.

46. Klieverik LP, Coomans CP, Endert E, et al. Thyroid hormone effects on whole-body energy homeostasis and tissue-specific fatty acid uptake in vivo. Endocrinology. 2009 Dec;150(12):5639–5648.

47. Santana-Farré R, Mirecki-Garrido M, Bocos C, et al. Influence of neonatal hypothyroidism on hepatic gene expression and lipid metabolism in adulthood. PLoS One. 2012;7(5):e37386.

48. Pascual G, Domínguez D, Elosúa-Bayes M, et al. Dietary palmitic acid promotes a prometastatic memory via Schwann cells. Nature. Nature Publishing Group; 2021 Nov;599(7885):485–490.

49. Oh DS, Lee HK. Autophagy protein ATG5 regulates CD36 expression and anti-tumor MHC class II antigen presentation in dendritic cells. Autophagy. Taylor & Francis; 2019 Dec 2;15(12):2091–2106.

50. Ruan C, Meng Y, Song H. CD36: an emerging therapeutic target for cancer and its molecular mechanisms. J Cancer Res Clin Oncol. 2022 Jul;148(7):1551–1558.

51. Rummel SK, Ellsworth RE. The role of the histoblood ABO group in cancer. Future Science OA [Internet]. Future Science; 2016 Jun [cited 2024 May 2];2(2). Available from: https://www.future-science.com/doi/10.4155/fsoa-2015-0012

52. Huang JY, Wang R, Gao Y-T, Yuan J-M. ABO blood type and the risk of cancer – Findings from the Shanghai Cohort Study. PLoS One. 2017 Sep 7;12(9):e0184295.

53. Zhan L, Chen L, Chen Z. Knockdown of FUT3 disrupts the proliferation, migration, tumorigenesis and TGF-β induced EMT in pancreatic cancer cells. Oncol Lett. 2018 Jul;16(1):924–930.

54. Nascimento JCF do, Beltrão EIC, Rocha CRC. High FUT3 expression is a marker of lower overall survival of breast cancer patients. Glycoconj J. 2020 Apr;37(2):263–275.

55. Lin L, Chen X, Lin G, Chen L, Xu Y, Zeng Y. FUT3 facilitates glucose metabolism of lung adenocarcinoma via activation of NF-κB pathway. BMC Pulmonary Medicine. 2023 Nov 9;23(1):436.

56. He C, Li A, Lai Q, et al. The DDX39B/FUT3/TGFβR-I axis promotes tumor metastasis and EMT in colorectal cancer. Cell Death Dis. Nature Publishing Group; 2021 Jan 12;12(1):1–12.

57. He M, Wu C, Xu J, et al. A genome wide association study of genetic loci that influence tumour biomarkers cancer antigen 19-9, carcinoembryonic antigen and α fetoprotein and their associations with cancer risk. Gut. 2014 Jan;63(1):143–151.

58. Holburn AM, Mach JP, MacDonald D, Newlands M. Studies of the association of the A, B and Lewis Blood group antigens with carcinoembryonic antigen (CEA). Immunology. 1974 Apr;26(4):831–843.

59. Yamashita K, Totani K, Kuroki M, Matsuoka Y, Ueda I, Kobata A. Structural studies of the carbohydrate moieties of carcinoembryonic antigens. Cancer Res. 1987 Jul 1;47(13):3451–3459.

60. Zhang T, Fassl A, Vaites LP, et al. Interrogating Kinase–Substrate Relationships with Proximity Labeling and Phosphorylation Enrichment. J Proteome Res. American Chemical Society; 2022 Feb 4;21(2):494–506.

61. Garnham R, Scott E, Livermore KE, Munkley J. ST6GAL1: A key player in cancer. Oncol Lett. 2019 Aug;18(2):983–989.

62. Anurag M, Strandgaard T, Kim SH, et al. Multiomics profiling of urothelial carcinoma in situ reveals CIS-specific gene signature and immune characteristics. iScience. 2024 Mar 15;27(3):109179.

63. Liu M, Jiang Y, Wedow R, et al. Association studies of up to 1.2 million individuals yield new insights into the genetic etiology of tobacco and alcohol use. Nat Genet. 2019 Feb;51(2):237–244.

64. Wu Y, Burch KS, Ganna A, Pajukanta P, Pasaniuc B, Sankararaman S. Fast estimation of genetic correlation for biobank-scale data. Am J Hum Genet. 2022 Jan 6;109(1):24–32.

65. Fry A, Littlejohns TJ, Sudlow C, et al. Comparison of Sociodemographic and Health-Related Characteristics of UK Biobank Participants With Those of the General Population. American Journal of Epidemiology. 2017 Nov 1;186(9):1026–1034.

66. Wang Y, Namba S, Lopera E, et al. Global Biobank analyses provide lessons for developing polygenic risk scores across diverse cohorts. Cell Genomics. 2023 Jan 11;3(1):100241.

67. Martin AR, Gignoux CR, Walters RK, et al. Human Demographic History Impacts Genetic Risk Prediction across Diverse Populations. Am J Hum Genet. 2017 Apr 6;100(4):635–649.

68. Lipsky JJ, Shen ML, Naylor S. In vivo inhibition of aldehyde dehydrogenase by disulfiram. Chem Biol Interact. 2001 Jan 30;130–132(1–3):93–102.

69. Shen ML, Johnson KL, Mays DC, Lipsky JJ, Naylor S. Determination of in vivo adducts of disulfiram with mitochondrial aldehyde dehydrogenase. Biochem Pharmacol. 2001 Mar 1;61(5):537–545.

70. Cho Y, Shin S-Y, Won S, Relton CL, Davey Smith G, Shin M-J. Alcohol intake and cardiovascular risk factors: A Mendelian randomisation study. Sci Rep. Nature Publishing Group; 2015 Dec 21;5(1):18422.

71. Jee YH, Lee SJ, Jung KJ, Jee SH. Alcohol Intake and Serum Glucose Levels from the Perspective of a Mendelian Randomization Design: The KCPS-II Biobank. PLoS One. 2016;11(9):e0162930.

72. Matoba N, Akiyama M, Ishigaki K, et al. GWAS of 165,084 Japanese individuals identified nine loci associated with dietary habits. Nat Hum Behav. 2020 Mar;4(3):308–316.

73. Yuan K, Longchamps RJ, Pardiñas AF, et al. Fine-mapping across diverse ancestries drives the discovery of putative causal variants underlying human complex traits and diseases [Internet]. medRxiv; 2023 [cited 2024 Jan 21]. p. 2023.01.07.23284293. Available from: https://www.medrxiv.org/content/10.1101/2023.01.07.23284293v4

74. Cai M, Wang Z, Xiao J, Hu X, Chen G, Yang C. XMAP: Cross-population fine-mapping by leveraging genetic diversity and accounting for confounding bias. Nat Commun. Nature Publishing Group; 2023 Oct 28;14(1):6870.

75. Bi W, Zhou W, Dey R, Mukherjee B, Sampson JN, Lee S. Efficient mixed model approach for large-scale genome-wide association studies of ordinal categorical phenotypes. The American Journal of Human Genetics. Elsevier; 2021 May 6;108(5):825–839.

76. Loh P-R, Tucker G, Bulik-Sullivan BK, et al. Efficient Bayesian mixed-model analysis increases association power in large cohorts. Nat Genet. Nature Publishing Group; 2015 Mar;47(3):284–290.

77. Mbatchou J, Barnard L, Backman J, et al. Computationally efficient whole-genome regression for quantitative and binary traits. Nat Genet. Nature Publishing Group; 2021 Jul;53(7):1097–1103.

78. Chang CC, Chow CC, Tellier LC, Vattikuti S, Purcell SM, Lee JJ. Second-generation PLINK: rising to the challenge of larger and richer datasets. GigaScience. 2015 Dec 1;4(1):s13742-015-0047–8.

79. Auton A, Abecasis GR, Altshuler DM, et al. A global reference for human genetic variation. Nature. Nature Publishing Group; 2015 Oct;526(7571):68–74.

80. Bulik-Sullivan B, Finucane HK, Anttila V, et al. An atlas of genetic correlations across human diseases and traits. Nat Genet. Nature Publishing Group; 2015 Nov;47(11):1236–1241.

81. Finucane HK, Bulik-Sullivan B, Gusev A, et al. Partitioning heritability by functional annotation using genome-wide association summary statistics. Nat Genet. Nature Publishing Group; 2015 Nov;47(11):1228– 1235.

82. Hujoel MLA, Gazal S, Loh P-R, Patterson N, Price AL. Liability threshold modeling of case-control status and family history of disease increases association power. Nat Genet. 2020 May;52(5):541–547.

83. Manichaikul A, Mychaleckyj JC, Rich SS, Daly K, Sale M, Chen W-M. Robust relationship inference in genome-wide association studies. Bioinformatics. 2010 Nov 15;26(22):2867–2873.

84. Mountjoy E, Schmidt EM, Carmona M, et al. An open approach to systematically prioritize causal variants and genes at all published human GWAS trait-associated loci. Nat Genet. Nature Publishing Group; 2021 Nov;53(11):1527–1533.

85. Verbanck M, Chen C-Y, Neale B, Do R. Detection of widespread horizontal pleiotropy in causal relationships inferred from Mendelian randomization between complex traits and diseases. Nat Genet. Nature Publishing Group; 2018 May;50(5):693–698.

86. Yengo L, Vedantam S, Marouli E, et al. A saturated map of common genetic variants associated with human height. Nature. 2022 Oct;610(7933):704–712.

87. Williams AT, Chen J, Coley K, et al. Genome-wide association study of thyroid-stimulating hormone highlights new genes, pathways and associations with thyroid disease. Nat Commun. Nature Publishing Group; 2023 Oct 23;14(1):6713.

88. Sterenborg RBTM, Steinbrenner I, Li Y, et al. Multi-trait analysis characterizes the genetics of thyroid function and identifies causal associations with clinical implications. Nat Commun. 2024 Jan 30;15:888.

89. Willer CJ, Li Y, Abecasis GR. METAL: fast and efficient meta-analysis of genomewide association scans. Bioinformatics. 2010 Sep 1;26(17):2190–2191.

90. Zeng J, Xue A, Jiang L, et al. Widespread signatures of natural selection across human complex traits and functional genomic categories. Nat Commun. 2021 Dec;12(1):1164.

91. Wen X, Stephens M. Using linear predictors to impute allele frequencies from summary or pooled genotype data. The Annals of Applied Statistics. Institute of Mathematical Statistics; 2010 Sep;4(3):1158–1182.

92. Brown BC, Asian Genetic Epidemiology Network Type 2 Diabetes Consortium, Ye CJ, Price AL, Zaitlen N. Transethnic Genetic-Correlation Estimates from Summary Statistics. Am J Hum Genet. 2016 Jul 7;99(1):76–88.

93. Wallace C. A more accurate method for colocalisation analysis allowing for multiple causal variants. PLOS Genetics. Public Library of Science; 2021 Sep 29;17(9):e1009440.

94. Pruim RJ, Welch RP, Sanna S, et al. LocusZoom: regional visualization of genome-wide association scan results. Bioinformatics. 2010 Sep 15;26(18):2336–2337.

